# Defects in lysosomal function and lipid metabolism in human microglia harboring a *TREM2* loss of function mutation

**DOI:** 10.1101/2022.07.05.22277068

**Authors:** Fabia Filipello, Shih Feng You, Sidhartha Mahali, Abhirami Kannan Iyer, Rita Martinez, Olena Korvatska, Wendy H Raskind, Bryan Bollman, Claudia Cantoni, Luca De Feo, Laura Ghezzi, Miguel Minaya, Anil G Cashikar, Jun-Ichi Satoh, Wandy Beatty, Marina Cella, Laura Piccio, Celeste M Karch

## Abstract

TREM2 is an innate immune receptor expressed by microglia in the adult brain. Genetic variation in the *TREM2* gene has been implicated in risk for Alzheimer’s disease and frontotemoral dementia, while homozygous *TREM2* mutations cause a rare leukodystrophy, Nasu-Hakola disease (NHD). Despite extensive investigation, the role of TREM2 in NHD pathogenesis remains poorly understood. Here, we investigate the mechanisms by which a homozygous stop-gain *TREM2* mutation (p.Q33X) contributes to NHD. Induced pluripotent stem cell (iPSC)-derived microglia (iMGLs) were generated from two siblings homozygous for the *TREM2* p.Q33X mutation (termed NHD), one related non-carrier, and one unrelated non-carrier. Transcriptomic analysis and biochemical assays revealed that iMGLs from NHD patients exhibited lysosomal dysfunction, downregulation of cholesterol metabolism genes, and reduced lipid droplets compared to controls. Also, NHD iMGLs displayed defective activation and HLA antigen presentation, which were restored by enhancing lysosomal biogenesis through mTOR-dependent and independent pathways. Alteration in lysosomal gene expression, such as decreased expression of genes implicated in lysosomal acidification (*ATP6AP2*) and chaperone mediated autophagy (*LAMP2*), together with reduction in lipid droplets were also observed in post-mortem brain tissues from NHD patients, thus closely recapitulating *in vivo* the phenotype observed in iMGLs *in vitro*. Our study provides the first cellular and molecular evidence that the *TREM2* p.Q33X mutation in microglia leads to a defect in lysosomal function. A better understanding of how microglial lipid metabolism and lysosomal machinery are altered in NHD and how these defects impact microglia activation may provide new insights into mechanisms underlying NHD and other neurodegenerative diseases.

## INTRODUCTION

Nasu-Hakola disease (NHD), also referred to as polycystic lipomembranous osteodysplasia with sclerosing leukoencephalopathy (PLOSL; OMIM 221770, OMIM 605086), is a rare autosomal recessive disorder characterized by a progressive presenile dementia and bone cysts. Approximately 200 NHD cases have been reported worldwide, primarily in Japan and Finland with few cases also reported in Italy [2, 7, 37, 40, 64, 77, 87].

Clinically, patients with NHD show recurrent pathological bone fractures, with frontal lobe syndrome and progressive dementia during the fourth decade of life [59, 72]. Pathologically, the brains of NHD patients exhibit extensive demyelination, astrogliosis, axonal loss, accumulation of axonal spheroids, calcification, and activation of microglia in the white matter of frontal and temporal lobes and the basal ganglia [3, 72]. Cortical deposition of amyloid beta (Aβ) and focal neocortical neurofibrillary pathology have been also described [31]. NHD is caused by homozygous pathogenic mutations in the genes *triggering receptor expressed on myeloid cells 2* (*TREM2*) or *TYRO protein tyrosine kinase binding protein* (*TYROBP*), alternatively named *DNAX-activation protein 12* (*DAP12*) [73, 74, 78]. Seventeen homozygous pathogenic mutations have been identified in *TREM2* or *DAP12* genes and are predicted to cause loss of function and lead to a similar disease phenotype [46, 48, 53, 95]. More recently, missense heterozygous variants in *TREM2* have been implicated in risk for late-onset Alzheimer’s disease [35, 38] and frontotemporal dementia [9, 36]. Thus, understanding the functional impact of *TREM2* gene variants has major implications for a number of dementias.

The TREM2 receptor is expressed in myeloid cell populations, including peripheral macrophages and microglia in the central nervous system (CNS) [43, 85]. TREM2 is a type I trans-membrane glycoprotein with an extracellular V-type immunoglobulin (Ig) ectodomain, a connecting stalk followed by a transmembrane region and a C-terminal tail [47]. At the cell surface, TREM2 receptor signaling occurs through association with the adaptor protein DAP12, which interacts with transmembrane domain of TREM2. While the ligand for TREM2 has not been identified, the TREM2 receptor has been shown to bind bacterial components (Daws et al., 2003); anionic and zwitterionic lipids (Wang et al., 2015); and myelin (Poliani et al., 2015). Activation of the TREM2/DAP12 complex mediates the recruitment of the protein tyrosine kinase Syk, resulting in the phosphorylation of downstream mediators such as PLC-γ, PI3K and Vav2/3, which lead to mTOR and mitogen-activated protein kinase (MAPK)-mediated downstream signaling [16, 79, 92]. These intracellular signals promote macrophage/microglia survival [70, 71, 93]; proliferation [71]; phagocytosis [88]; synapse elimination [27]; and cellular metabolism [92]. The TREM2 receptor can also be cleaved by metalloproteases to produce a soluble form (sTREM2) which is detected in the cerebrospinal fluid (CSF) [26, 80]. Some of the NHD-causing missense mutations in *TREM2* likely affect TREM2 protein folding and stability, but the cellular mechanisms and the pathways dysregulated during the disease are still unknown [49]. Despite the predicted loss of function effect of NHD mutations in *TREM2*, mouse models of *Trem2* deficiency fail to capture key pathological aspects of NHD. As such, there remains a critical gap in our understanding of the pathologic impact of NHD mutations in the CNS and our ability to develop novel therapeutics.

Human stem cell models have emerged as a powerful cellular system that enables modeling of rare mutations in the cell-types affected in disease, including those that cause NHD [12, 18, 30, 68]. Yet, the extent to which stem cell models capture disease relevant phenotypes remains uncertain. To begin to define the impact of a rare loss of function mutation in *TREM2* on microglia function, we generated macrophages and human induced pluripotent stem cell (iPSC)-derived microglia like cells (iMGLs) from a family affected by NHD. The two affected siblings in the family are homozygous for the *TREM2 p.*Q33X mutation and their healthy sibling is homozygous for the wild-type allele (*TREM2* WT). *TREM2* p.Q33X introduces an early stop codon and is subject to nonsense mediated decay; hence, there is no TREM2 protein product [75]. Herein, transcription profiling and functional analyzes of the *TREM2* p.Q33X mutation reveal that it leads to dysregulation of lysosomal function, lipid metabolism and microglia activation, and these phenotypes are recapitulated in brain tissues from NHD patients.

## RESULTS

### Generation of iMGLs from control and NHD donors

To understand the contribution of a *TREM*2 mutation to myeloid cell dysfunction in NHD, dermal fibroblasts were obtained from a family consisting of two siblings affected by NHD and homozygous for the *TREM2 p.Q33X* mutation (referred to as NHD1 and NHD2) and a third sibling who is homozygous for *TREM2* WT (referred to as SB CTRL; Fig. 1A, B and Table S1). A second, unrelated control was also included in this study [41] (referred to as NR CTRL). Fibroblasts were reprogrammed into iPSC and characterized for pluripotency and genomic stability (Fig. S1). Blood samples from the three siblings were collected and peripheral blood mononuclear cells (PBMCs) were isolated to generate macrophages, a type of peripheral myeloid cells (Fig. 1C).

**Figure 1:**
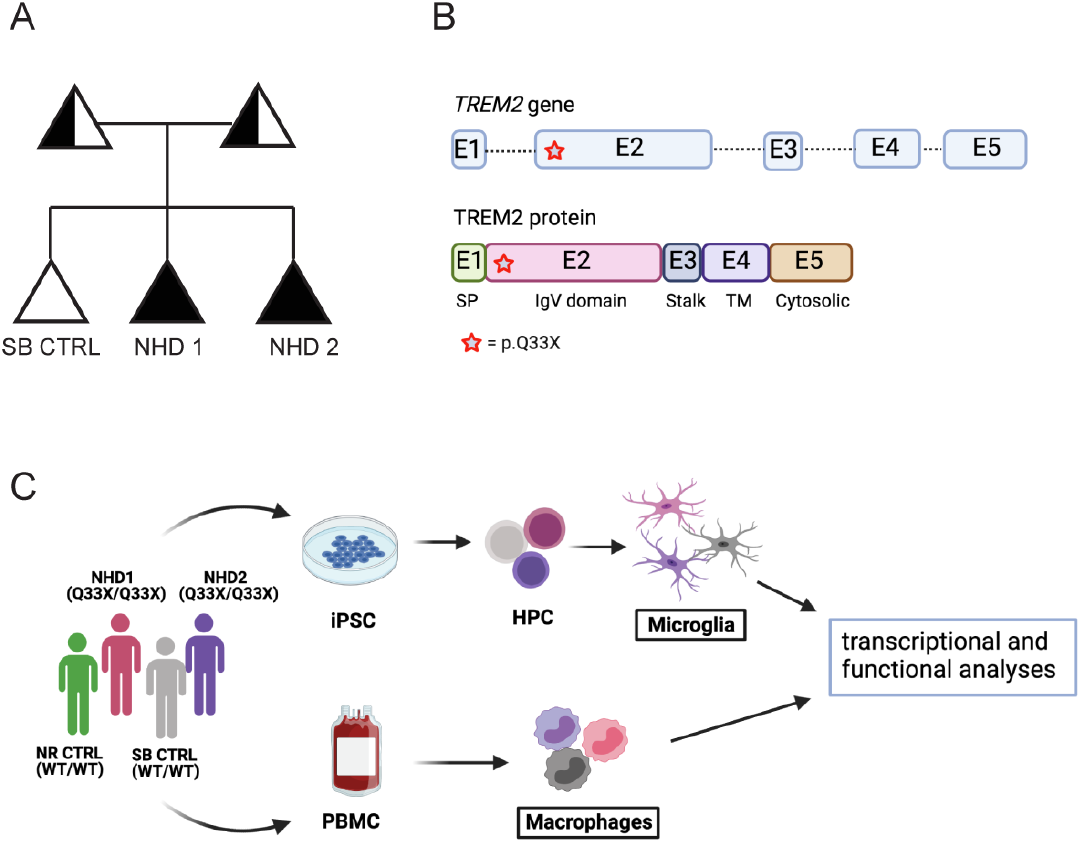
*TREM2* p.Q33X variant in a family affected by NHD. A) Family pedigree. Black symbols denote individuals clinically affected by NHD who carry the homozygous *Q33X* mutation in *TREM2* (NHD1 and NHD2). White symbols denote a clinically healthy, non-carrier family member (SB CTRL). Half shaded symbols denote heterozygous *TREM2 p.Q33X* carriers. Analyses presented in this manuscript were performed on the three siblings (SB CTRL, NHD1 and NHD2) and one unrelated control (NR CTRL). Triangles are used to anonymize pedigree. B) Schematic representation of the human *TREM2* gene (top) and protein (bottom). *TREM2* gene showing exonic (numbered boxes) and intronic (line) regions. The *Q33X* mutation is located in exon 2 (indicated as a red star). C) Study design depicting the bioinformatic and cellular *in vitro* analyses performed in this study. iPSC-derived human microglia (iMGLs) were generated from iPSCs, while human macrophages were purified from peripheral blood mononuclear cells (PBMCs)-derived from the same donors. Fully differentiated iMGLs and macrophages were assayed for molecular and biochemical analyses.

Microglia are the resident macrophages of the brain and the primary cell-type of myeloid origin in the CNS that expresses TREM2. To define the impact of *TREM2* p.Q33X on microglia gene expression and function, we generated iMGLs using a two-step differentiation protocol as previously described [1, 62] (Fig. 1C and Fig. S2A). This method generates iMGLs that are positive for the microglial markers TMEM119, P2RY12, IBA1, and TREM2 (Fig. S2B). iMGLs release sTREM2 in the media, which increases over time in culture and peaks when iMGLs are fully mature (Fig. S2C). iMGLs are distinct from peripheral macrophages derived from PBMCs and express intermediate CD45 (CD45^int^) levels compared to higher levels (CD45^high^) observed in macrophages (Fig. S2D). Additionally, iMGLs produce a gene expression signature that is distinct from macrophages based on principal component analysis of transcriptomic data (Fig. S2E). iMGLs are also enriched for microglia-specific genes including *CX3CR1*, *TGFBR1*, *P2RY12*, *GPR34* and *IRF8* [29, 97] compared with macrophages (fold change > 20; Fig. S2F and Table S2). Thus, the microglia differentiation protocol allows us to generate a functionally distinct myeloid population similar to CNS microglia, consistent with previous reports [1, 62].

We next sought to determine the impact of *TREM2* p.Q33X on microglia identity. iMGLs were differentiated from the NHD cases (NHD1 and NHD2) and the healthy sibling control (SB CTRL) (Fig. 2A). Transcriptomic analyses revealed that microglia-enriched genes are similarly expressed in the SB CTRL, NHD1 and NHD2 iMGLs (Fig. S2F). Thus, NHD and control iMGLs maintain a similar propensity to form fully differentiated microglia-like cells. As expected, *TREM2* mRNA transcripts were significantly reduced in NHD1 and NHD2 iMGLs compared to NR and SB CTRL (p=4.28×10^−41^, Fig. S2F and Fig. 2B), while *TYROBP* mRNA levels were comparable across iMGLs (Fig. 2C). TREM2 protein was present at the membrane and in the intracellular compartment in SB CTRL iMGLs but was absent in NHD1 and NHD2 iMGLs by flow cytometry (FACS) (Fig. 2D-F) and immunofluorescence (Fig. 2G). Additionally, SB CTRL iMGLs produced robust sTREM2 levels in culture medium while sTREM2 was absent in medium from NHD1 and NHD2 iMGLs (Fig. 2H). Together, these findings illustrate that the *TREM2* p.Q33X mutation negatively impacts the production of a functional membrane TREM2 and soluble TREM2 receptor in iMGLs.

**Figure 2:**
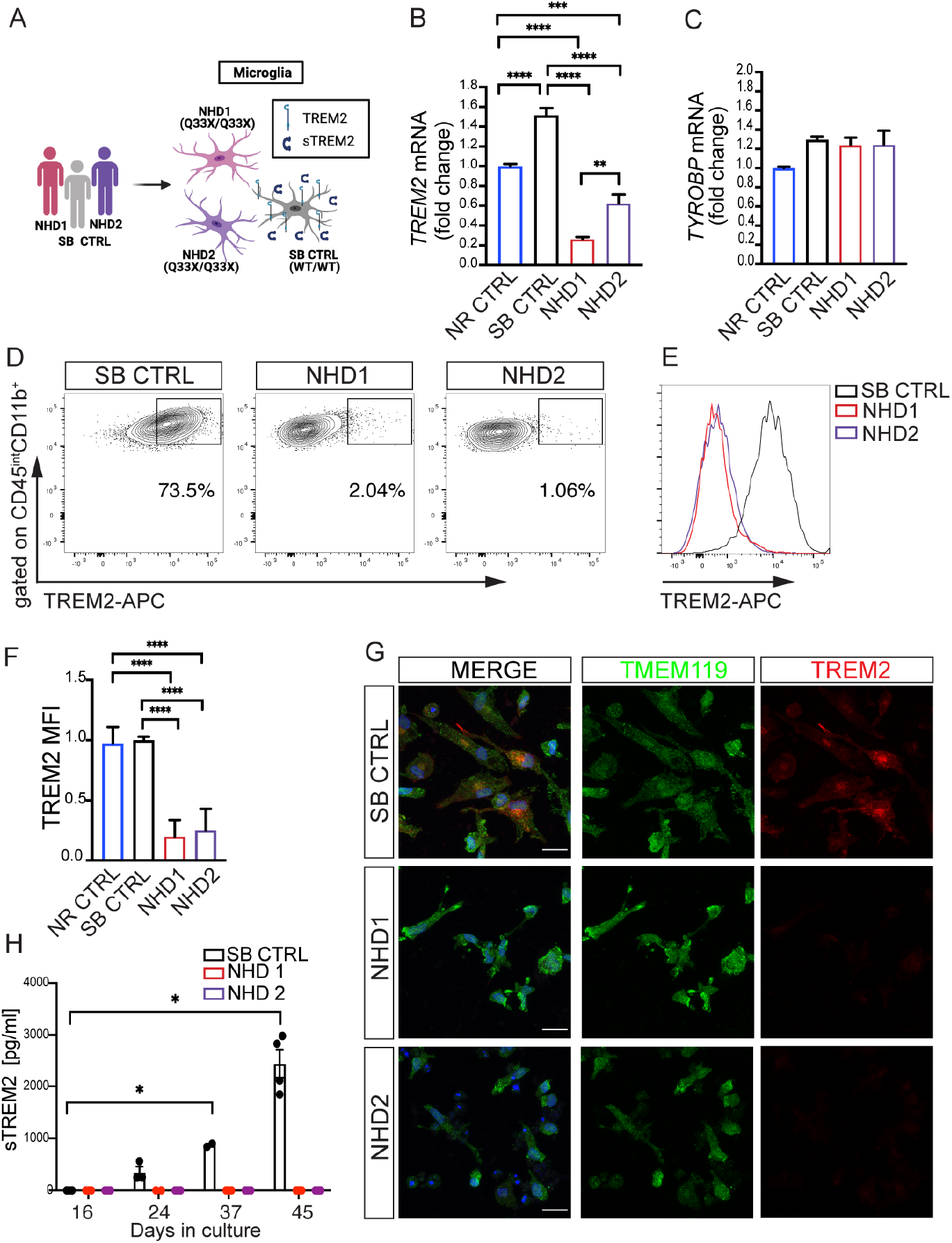
*TREM2* p.Q33X mutation abolishes cell surface expression of TREM2 receptor on iMGLs and abrogates sTREM2 production. A) Diagram representing patient-derived iMGLs. B) and C) RT-qPCR of *TREM2* and *TYROBP* mRNA in NR CTRL, SB CTRL, NHD1 and NHD2 iMGLs. ***P < 0.001, ****P < 0.0001, One-way ANOVA with Tukey’s post hoc test. Data are pooled from two independent experiments. D-F) FACS analysis of TREM2 receptor expression at the surface of iMGLs. D) Representative dot plots showing the percentage of TREM2-positive cells in the CD45^int^ CD11b^+^ population. E) Representative histogram of TREM2 mean fluorescent intensity (MFI) in SB CTRL, NHD1 and NHD2 iMGLs. F) Relative quantification of TREM2 MFI in NR CTRL, SB CTRL, NHD1 and NHD2 iMGLs. ****P < 0.0001, One-way ANOVA with Tukey’s post hoc test. Data are pooled from three independent experiments. G) Representative confocal images of iMGLs from SB CTRL, NHD1 and NHD2 stained for TMEM119 (green), TREM2 (red) and DAPI (blue). Scale bar 10 μm. H) Media from SB CTRL, NHD1 and NHD2 iMGLs were collected at different times during iMGLs differentiation process and sTREM2 was measured by ELISA. *P < 0.05, Two-way ANOVA with Dunnett’s post hoc test. Data are pooled from two independent experiments. Data shown are mean ± SEM.

Beyond direct effects on TREM2, the *CCL3* gene (macrophage inflammatory protein-1 a, MIP-1a) was significantly reduced in NHD iMGLs compared to SB CTRL (p=2.42×10^−24^, Fig. S2F). *CCL3* is a neutrophil chemoattractant and a direct stimulator of osteoclastogenesis, a process that together with bone remodeling is controlled by TREM2/DAP12 signaling and is altered in NHD patients [10, 59, 65, 70]. We also observed a higher number of annexin-V^+^ (AnxV) propidium iodide (PI)^+^ cells in both NHD1 and NHD2 iMGLs compared to controls (Fig. S2G and H), with NHD2 showing more cell death than NHD1 (Fig. S2H). This finding is consistent with a role for TREM2 in sustaining microglial survival [63, 71, 93] and suggests that we can capture disease-relevant signatures in a dish.

### Downregulation of antigen presentation and immune activation pathways in *NHD* iMGLs and macrophages

To understand the broad impact of *TREM2* p.Q33X on microglia function, we performed transcriptomic analyses in iMGLs generated from SB CTRL, NHD1 and NHD2. We identified 3353 differentially expressed genes when comparing NHD iMGLs with SB CTRL (using a cutoff of ±1 log fold change and an FDR < 0.05; Table S2 and Fig. S3A and B). We found that the genes upregulated in NHD iMGLs (1950 genes) belong to pathways associated with CNS demyelination (p=9.36×10^−4^), leukodystrophies (p=0.02), and bone pain (p=0.04) (Fig. 3A and Table S3). Among the genes downregulated in NHD iMGLs (1816 genes), we observed an enrichment in genes involved in NF-κB signaling (p=1.46×10^−11^), osteoclast differentiation (p=2.06×10^−8^), tumor necrosis factor (TNF) signaling (p=1.58×10^−7^), phagosome (p=3.76×10^−5^), lysosome (p=7.31×10^−5^), and Toll-like receptor (TLR) signaling (p=1.37×10^−4^) (Fig. 3A and Table S3). Genes involved in human leukocyte antigen (HLA) presentation and belonging to the major histocompatibility complex (MHC) class I and MHC class II clusters were also significantly reduced in NHD iMGLs compared to SB CTRL: *HLA.F, TAPBP, HLA.B, CALR, HLA.E, TAP2, TAP1, HLA.DPA1, HLA.DRB1, HLA.DRA, HLA.DOB, HLA.DQA1, CD74* (Fig. 3B). The molecular findings of a defect in immune activation in NHD iMGLs were confirmed at the protein level by flow cytometry, which demonstrated a significant reduction of the costimulatory molecules such as CD80 and CD86 (Fig. 3C-E), HLA-DR (Fig. 3C, 3F, 3G and 3H), and CD14 (Fig. 3I) at the cell surface of NHD iMGLs compared to controls. A decrease of immunostaining of the phagocytic marker CD68 in NHD iMGLs was also observed, consistent with an overall decrease in activation (Fig. 3J and K). Thus, under basal conditions, NHD iMGLs express a molecular and cellular phenotype consistent with a hypo-reactive phenotype.

**Figure 3:**
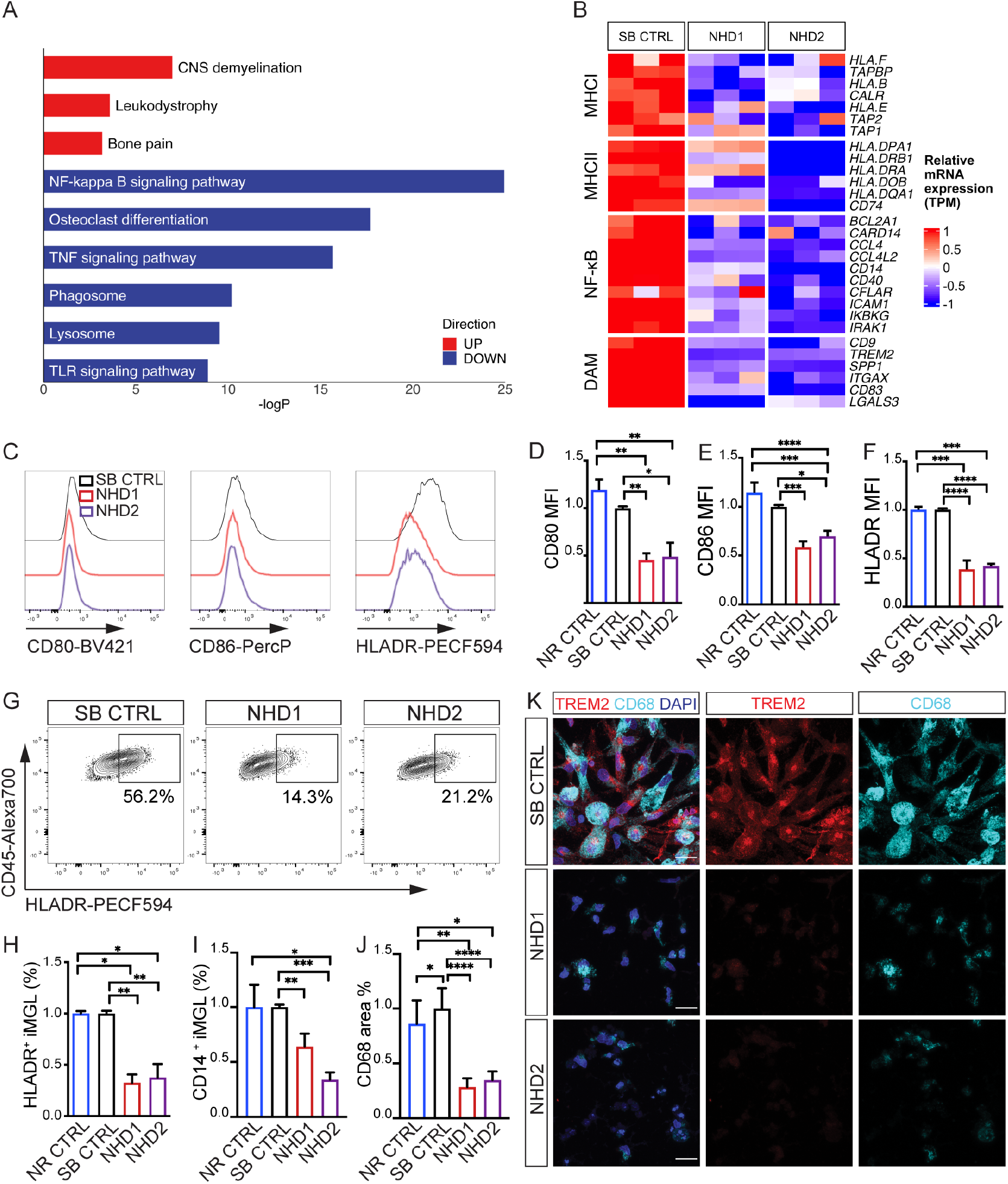
Defective immune activation of NHD iMGLs. A) Pathway analysis of protein coding differentially expressed genes (DEG) in NHD *versus* SB CTRL iMGLs (EnrichR; DEG are defined by FDR < 0.05, DEGs in Table S2). B) Heatmap of DEG belonging to MHC I, MHC II, NF-κB and DAM clusters in iMGLs from SB CTRL *versus* NHD subjects (N=1 SB CTRL, N=2 NHD lines - NHD1 and NHD2-; n=3 replicates/line). C) Representative histograms and D-F) relative quantification performed by FACS showing cell surface D) CD80, E) CD86 and F) HLA-DR MFIs in NR CTRL, SB CTRL, NHD1 and NHD2 iMGLs. G) Representative dot plots showing the percentage of HLADR-positive cells in SB CTRL, NHD1 and NHD2 iMGLs. H) Relative quantification of HLADR^+^ and I) CD14^+^ cells percentage in NR CTRL, SB CTRL, NHD1 and NHD2 iMGLs. *P < 0.5, **P < 0.01, ***P < 0.001, ****P < 0.0001, One-way ANOVA with Tukey’s post hoc test. Data are pooled from four independent experiments. J) Quantification of CD68 area % in NR CTRL, SB CTRL, NHD1 and NHD2 iMGLs and K) representative confocal images of iMGLs stained for CD68 (cyan), TREM2 (red) and DAPI (blue). *P < 0.05, **P < 0.01, ****P < 0.0001, One-way ANOVA with Kruskal Wallis test and Dunn’s multiple comparison test. Data are pooled from two independent experiments. Data shown are mean ± SEM.

To determine whether the effects of *TREM2* p.Q33X are conserved across myeloid cell populations, we compared transcriptomic data generated from PBMC-derived macrophages from the three siblings along with the iMGLs described above (Fig. S3A). NHD macrophages produced a significant dysregulation of genes (2266 genes) enriched in pathways associated with proteasome (p=1.58×10^−11^), antigen processing and presentation (p=7.52×10^−6^), and steroid biosynthesis (p=2.27×10^−7^) compared to SB CTRL (Fig. S3C and D and Table S4). We identified 345 genes that were commonly differentially expressed between NHD and SB CTRL in iMGLs and macrophages (Fig. S3E and F). These commonly differentially expressed genes were enriched in pathways involved in phagocytosis (p=3.38×10^−4^), response to interferon gamma (IFNγ) (p=1.40×10^−3^), antigen processing and presentation (p=1.23×10^−2^), regulation of apoptotic signaling pathways (p=3.94×10^−3^), and lipid catabolic process (p=9.36×10^−3^) (Fig. S3G). Thus, *TREM2* p.Q33X results in the dysregulation of phagocytic, activation, and lipid metabolic pathways across the myeloid lineage.

Trem2 expression is required for the transition to disease associated microglia (DAM) in mouse models of neurodegeneration [42, 51] and in human microglia transplanted in a mouse model of amyloid accumulation [63]. Thus, we sought to determine the impact of *TREM2* p.Q33X on genes enriched in the DAM signature in NHD and SB CTRL iMGLs. We found that the DAM genes were reduced in NHD iMGLs (Fig. 3B), consistent with the requirement of *Trem2* for these signatures (Fig. 3B) [42]. *Trem2* deficiency is associated with an overactive response to bacterial lipopolysaccharide (LPS)-induced inflammation in mouse bone marrow-derived macrophages [57, 91], but studies using human iMGLs from NHD mutation carriers have generated conflicting observations [12, 30]. We investigated the effect of LPS, a well-characterized activator of TLR4 and a binding partner of TREM2 [19] in control and NHD iMGLs. We found that LPS treatment led to significantly elevated levels of HLA-DR, CD80, and CD86 in NHD iMGLs compared to control iMGLs, suggesting an exaggerated response (Fig. S4A-C). Thus, NHD iMGLs exhibit hypoactivation under basal conditions and hyperactivation in response to inflammatory stimuli.

### Lysosomal defects in NHD iMGL

*Trem2* deficient mice fail to upregulate genes involved in lysosomal and degradative pathways during demyelination [67, 92], suggesting a role for TREM2 in endolysosomal function. However, the impact of TREM2 mutations on endolysosomal function has not been fully explored. We found that genes involved in endolysosomal pathways (*LAMP1*, *LAMP2*, *RAB7A*, *CTSS*, *CD63*, *CD68*, and *RAB5C*) were significantly downregulated in NHD compared with SB CTRL iMGLs (Fig. 3A and Fig. 4A). In order to determine whether these gene signatures are driven by the loss of function effect of the *TREM2* p.Q33X, we examined transcriptomic data from *TREM2* WT and CRISPR/Cas9-engineered *TREM2* knockout (KO) iMGLs [63]. The absence of *TREM2* led to the reduction of 2297 genes in the iMGL isogenic pairs compared with the 1816 genes downregulated in NHD *versus* SB CTRL iMGLs, of which 268 genes were shared (Fig. 4B and Table S5). Pathway analysis of the 268 commonly downregulated genes revealed an enrichment in TLRs (*TRAF3, CCL3L3, CCL4, CCL3, SPP1, LY96*); lysosome (*NPC2, HEXB, LAMP2, AP1B1, CD68, CTSD, CTSC*); and cholesterol metabolism (*CYP27A1, SOAT1, NCP2, LPL*; Fig. 4B). Thus, loss of TREM2 function (by gene mutation or deletion) drives altered immune response, lysosome function and cholesterol metabolism.

**Figure 4:**
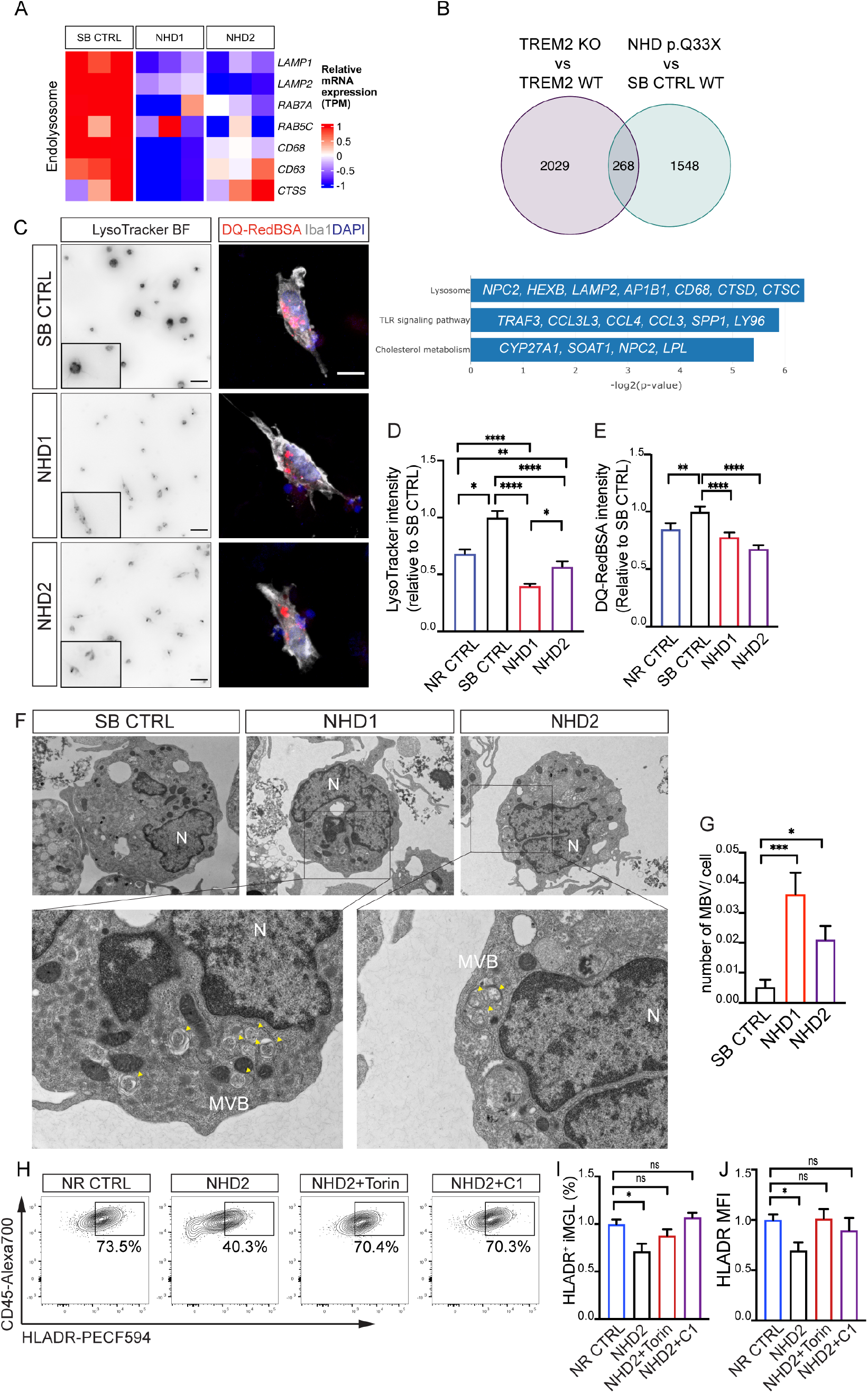
Lysosomal defects and MVBs accumulation in NHD iMGLs. A) Heatmap of the endo-lysosomal genes *LAMP1, LAMP2, RAB7A, CTSS, CD63, CD68, RAB5C* in iMGLs from SB CTRL versus NHD subjects (N=1 SB CTRL, N=2 NHD lines; n=3 replicates/line). B) Venn diagram of downregulated genes in common between CRISPR/Cas9-engineered *TREM2* knockout (KO) *versus* WT published in [63] and NHD *versus* SB CTRL (p<0.05; genes in common=268) and pathway analysis of the 268 downregulated genes in common between TREM2 KO *versus* WT and NHD *versus* SB CTRL iMGLs. C) Representative bright field images and insets showing iMGLs stained with LysoTracker (left); representative confocal images of iMGLs stained with DQ-Red-BSA (red), Iba1 (grey) and DAPI (blue) (right). Scale bar 10 μm. Relative quantification of D) LysoTracker and E) DQ-Red-BSA intensity in NR CTRL, SB CTRL, NHD1 and NHD2 iMGLs. LysoTracker: NR CTRL n= 127 cells; SB CTRL= 156 cells, NHD1 n=168 cells; NHD2 n=107 cells. DQ-Red-BSA: NR CTRL n= 131 cells; SB CTRL= 141 cells, NHD1 n=177 cells; NHD2 n=145 cells. *P < 0.05, **P < 0.01, ****P < 0.0001, One-way ANOVA with Kruskal Wallis test and Dunn’s multiple comparison test. Data are pooled from two independent experiments. F) Representative electron microscopy images of SB CTRL, NHD1 and NHD2 iMGLs and higher magnification. Undigested material (lamellar structures) is detected within multivesicular bodies (MVBs) in NHD1 and NHD2 iMGLs (yellow arrowheads). N, nucleus. Scale bar 2 μm (top) and 500 nm (bottom). G) Quantification of the number of late endosome/MVBs with inclusion bodies normalized per cell area in SB CTRL, NHD1 and NHD2 iMGLs. SB CTRL n= 18 cells; NHD1 n= 18 cells; NHD2 n= 17 cells. *P < 0.05, ***P < 0.001, One-way ANOVA with Kruskal Wallis test and Dunn’s multiple comparison test. H) Representative dot plots showing the percentage of HLA-DR-positive cells in NR CTRL, NHD2, NHD2 + Torin1 (250nM), NHD2 + curcumin analog C1 (1 μM) iMGLs. I) Relative quantification of HLA-DR^+^ cells percentage and J) HLA-DR MFI. *P < 0.05, analysis was performed between two groups using an Unpaired T test. Data are pooled from two independent experiments. Data shown are mean ± SEM.

To determine whether the impact of *TREM2* p.Q33X on lysosomal gene expression leads to functional defects in lysosomal machinery, we measured lysosomal acidification in NHD, NR and SB CTRL iMGLs using the pH-sensitive dye, LysoTracker red DND-99 (Fig. 4C). LysoTracker fluorescence was significantly reduced in NHD iMGLs compared to control iMGLs (Fig. 4C and D), indicating defective vesicle acidification. To determine whether *TREM2* p.Q33X also affects proteolytic degradation, we treated NHD and control iMGLs with DQ-Red BSA, which captures cargo delivery and degradation [61]. DQ-Red BSA intensity was significantly reduced in NHD1 and NHD2 iMGLs compared to control iMGLs, suggesting a defect in uptake or breakdown of the substrate (Fig. 4C and E). Additionally, using electron microscopy, we detected an accumulation of unprocessed material within multivesicular bodies (MVBs) [33] in NHD1 and NHD2 iMGLs compared to the SB CTRL (Fig. 4F and 4G). These findings suggest that *TREM2* p.Q33X, likely via TREM2 functional deficiency, leads to defects in lysosomal function that result in the accumulation of undegraded material in MVBs (Fig. S4D).

Proper assembly and recycling of MHC class II molecules to the cell membrane requires the endolysosomal system [17, 28]. As such, our observation of reduced HLA-DR expression in NHD iMGLs may be a consequence of lysosomal dysfunction. In addition to the core lysosomal genes that were downregulated in NHD iMGLs, we observed a significant reduction of *TFEB* (p=1.1×10^−2^) (Table S2). *TFEB* is the master regulator of lysosomal biogenesis and positively controls autophagosome formation and autophagosome-lysosome fusion [84, 86]. To determine whether restoring lysosomal function could rescue key defects in the NHD iMGLs, we tested the impact of compounds that activate TFEB via mTOR-dependent (Torin) and -independent (curcumin c1) pathways. After pre-treatment with Torin or curcumin c1, we fully rescued HLA-DR expression in NHD iMGLs (expressed as MFI and cell percentage) to levels comparable to control iMGLs (Fig. 4H-J). This finding suggests that rescuing lysosomal abnormalities in NHD iMGLs may have broad restorative effects.

### Lipid metabolism is reduced in NHD iMGL

Lysosomal pathways play a critical role in processing and sorting exogenous and endogenous lipids [90]. *Trem2* is required for lipid droplet biogenesis in a toxin-induced model of CNS demyelination [32], and *Trem2*–deficient mice exhibit defects in lipid droplet formation due to excessive cholesterol [32]. *TREM2* p.Q33X iMGLs resulted in a downregulation of cholesterol pathway genes (*CYP27A1, SOAT1, NCP2, LPL*) (Fig. 4B and Fig. 5A). Thus, we sought to investigate whether NHD iMGLs display defects in lipid droplet biogenesis. Electron microscopy revealed a significant decrease of lipid droplet number in NHD2 compared to SB CTRL iMGLs (Fig. 5B and C). NHD1 showed a reduction of lipid droplet size compared to SB CTRL iMGLs without a significant change in droplet numbers (Fig. 5B-D). Additionally, we observed a significant reduction in the intensity of Perlipin2 (PLIN2) expression, a protein that coats intracellular lipid droplets, in NHD compared to NR and SB CTRL iMGLs (Fig. 5E and F). Staining iMGLs with the fluorescent neutral lipid dye Bodipy, which illuminates neutral lipids, revealed a decrease in Bodipy intensity in NHD1 and NHD2 iMGLs compared to SB CTRL (Fig. S4E and F). Myelin debris are actively phagocytosed by microglia during demyelination to promote clearance and repair of the brain, with TREM2 receptor playing a pivotal role in this process [15, 82]. We observed that control and NHD iMGLs were able to engulf pHrodo-conjugated human myelin, as shown by confocal acquisition (Fig. 5G and H). To investigate whether cholesterol-rich myelin has an effect on lipid droplet content in NHD iMGL, we next treated control and NHD iMGLs with human myelin for 24h. Even in the presence of myelin, we still detected a reduced amount of PLIN2^+^ staining in NHD iMGL compared to NR and SB CTRL (Fig. 5I and J). Overall, these results show that NHD iMGLs display impaired lipid droplet formation and cholesterol metabolism both in the presence and absence of myelin.

**Figure 5:**
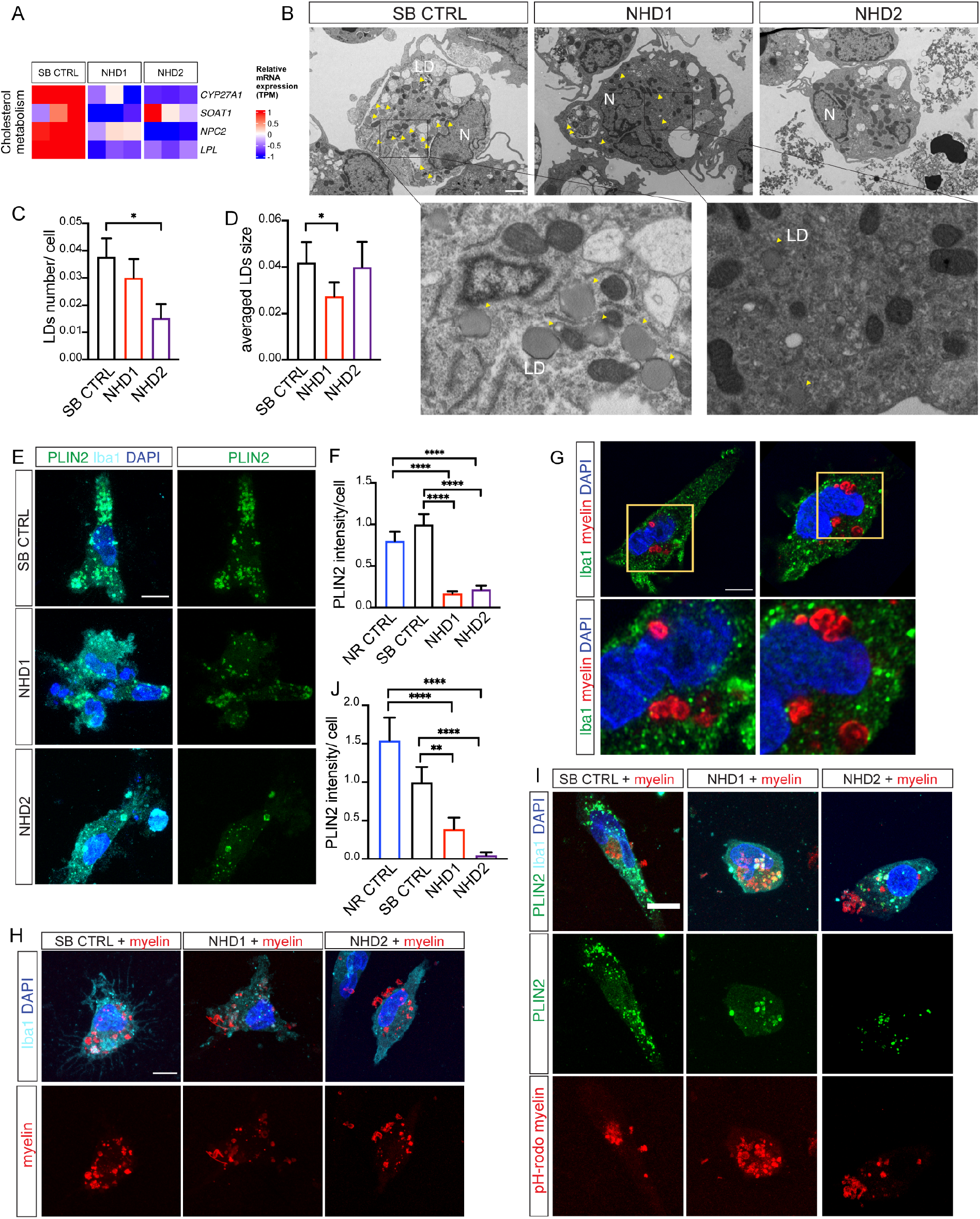
iMGLs from NHD patients display reduced number of lipid droplets. A) Heatmap of *CYP27A1, SOAT1, NCP2, LPL* genes in iMGLs from SB CTRL versus NHD subjects (N=1 SB CTRL, N=2 NHD lines -NHD1 and NHD2-; n=3 replicates/line). B) Representative electron microscopy images of SB CTRL, NHD1 and NHD2 iMGLs showing lipid droplets (LD) (yellow arrowheads). At the bottom are enlarged images of the enclosed area. N, nucleus. Scale bar 2mm. C) Quantification of the number of lipid droplets per cell area in SB CTRL, NHD1 and NHD2 iMGLs. D) Quantification of lipid droplets size in SB CTRL; NHD1 and NHD2 iMGL. SB CTRL n= 18 cells; NHD1 n= 18 cells; NHD2 n= 17 cells. *P < 0.05, One-way ANOVA with Tukey’s post hoc test. E) Representative confocal images of Perilipin2 (PLIN2) (green) in Iba-1^+^ iMGLs (cyan), DAPI (blue). Scale bar 10μm. F) PLIN2 intensity quantification. NR CTRL n= 77 cells; SB CTRL= 101 cells, NHD1 n=97 cells; NHD2 n=69 cells. ****P < 0.0001, One-way ANOVA with Kruskal Wallis test and Dunn’s multiple comparison test. Data are pooled from three independent experiments. G) Representative confocal images of SB CTRL stained with Iba1 (green) and treated with pH-rodo human myelin (red) for 24 h. At the bottom are enlarged images of the enclosed area. Scale bar 10μm. H) Representative confocal images of SB CTRL, NHD1 and NHD2 iMGLs stained with Iba1 (cyan) and DAPI (blue) and engulfing pH-rodo human myelin (red) (24h treatment). I) Representative confocal images of PLIN2 (green) in Iba-1^+^ iMGLs (cyan), DAPI (blue), treated with pH-rodo human myelin (red) for 24h. Scale bar 10μm. J) PLIN2 intensity quantification. NR CTRL n= 37 cells; SB CTRL= 39 cells, NHD1 n=47 cells; NHD2 n=39 cells. **P < 0.01, ****P < 0.0001, One-way ANOVA with Kruskal Wallis test and Dunn’s multiple comparison test. Data are pooled from two independent experiments. Data shown are mean ± SEM.

### Lysosomal and metabolic defects in NHD brain tissues

To determine the extent to which the NHD iMGLs phenotypes are represented in NHD patients, we examined brain tissues from NHD and healthy controls. First, we analyzed gene expression data by Nanostring in hippocampi [50] from four NHD patients and 17 healthy controls (Table S1). We identified 122 significantly upregulated genes and 46 downregulated genes among the 800 genes evaluated (p value < 0.05) (Fig. 6A and Table S6). Genes involved in neuroplasticity, synaptic function (*C1QBP, APP*, *NEFL*, *LRRN3*, *MEF2C*, *THY1*), and neuron-microglia crosstalk (*CD200* and *IL34*) were downregulated in NHD brains (Fig. 6A and Table S6). We also observed a significant increase in genes associated with microgliosis (*CX3CR1* and *AIF*; p=1.58×10^−3^ and p=0.04, respectively) and astrogliosis (*S100B*; p=4.22×10^−4^). The genes significantly upregulated in NHD brains were enriched in pathways associated with inflammation and immune activation (Fig. 6A), such as inflammatory cytokines (*IL8, IL18, IL6R, TGFB1, IL17RA)*; antigen presentation molecules (*IRF8, CD86*, *HLA-DPB1, HLA-DRA*); and chemokines (*CXCL2, CXCL12, CXCL16, SELPLG*) (Table S6). These findings illustrate that post-mortem brain tissues from NHD patients display a profound effect on immune cell activation and inflammation, which is consistent with the dysregulation of these pathways in iMGLs and with the exaggerated response to LPS. Among the downregulated genes in NHD brains, we observed genes enriched in pathways involved in the activation of mitogen-activated protein kinase (MAPK)-mediated cascade and NF-κB (*PRKCE, DUSP4, MAP2K1, MAP2K, AKT3, ECSIT)* [79, 95]. Strikingly, *ATP6AP2* and *LAMP2,* which are involved in lysosomal acidification and lysosomal protein degradation [23, 44], were significantly downregulated in NHD brains compared to controls (p=1.44×10^−4^ and p=5.95×10^−3^, respectively) (Fig. 6B and C). This finding supports our observations of lysosomal functional defects in NHD iMGLs (Fig. 4). Thus, we showed that key phenotypic features observed in *TREM2* p.Q33X iMGLs, such as defects in immune activation and lysosomal dysfunction, are conserved in human brains from NHD patients.

**Figure 6:**
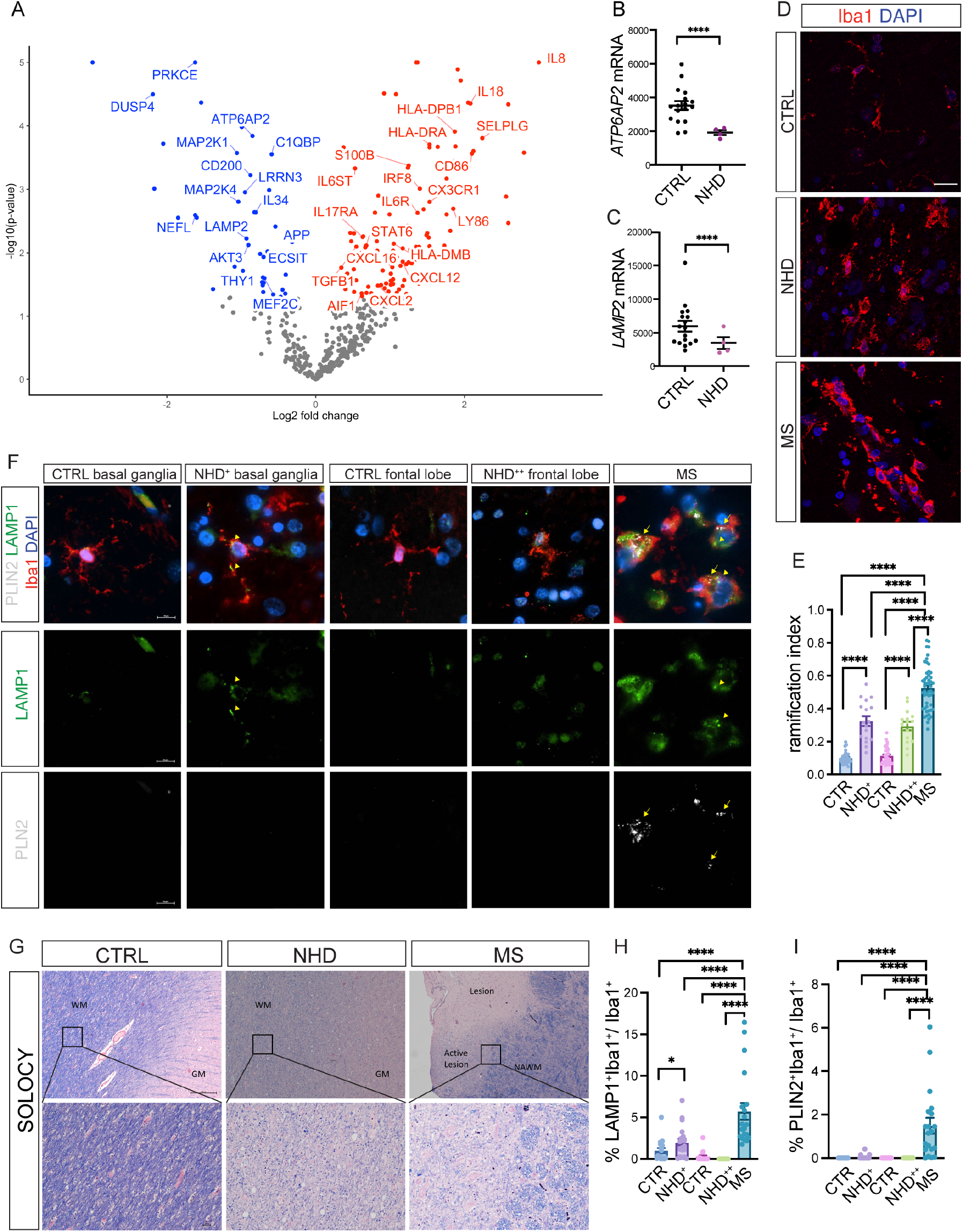
Post-mortem tissues from NHD patients display lysosomal and lipid metabolism defects. A) NanoString analysis was performed on hippocampi from four NHD patients (one *TREM2 D134G*; one *TREM2* c.482+2T>C; two *TYROBP* c.141delG) and 17 controls. Volcano plot of DEG between control and NHD brains (p < 0.05). Red: upregulated genes; blue: downregulated genes. B) *ATP6AP2*, and C) *LAMP2* mRNA (raw counts) from NanoString analysis of NHD *versus* control brains. ****P<0.0001. D) Representative confocal images of Iba1^+^ microglia (red) and DAPI (blue) staining of frontal lobe (FL) regions of a healthy control, one NHD patient (NHD1^++^ (FL)), and one MS patients (active lesion in the medulla). Scale bar 20μm. E) Microglia morphometric analysis in the basal ganglia (BG) and frontal lobe (FL) regions of three controls, two NHD patient (NHD1^+^ (BG), NHD1^++^ (FL)), and two MS patients (active lesion region). The analysis was done using a ramification index [RI = 4π × cell area/(cell perimeter)^2^] that describes microglia cell shape. BG: CTRL N=3, n= 31 cells; NHD1^+^ N=1, n= 17; FL: CTRL N=3, n= 38 cells; NHD1^++^ N=1, n= 15; MS N=2, n= 50. ****P<0.0001, analysis was performed between two groups using a Mann-Whitney T test. F) Representative images of Iba1^+^ microglia (red), PLIN2 (grey), LAMP1 (green) and DAPI (blue) staining of FL and BG of controls, NHD patients and one MS patient (active lesion in the medulla). Scale bar 20μm. G) White matter (WM) and grey matter (GW) regions with no pathology in one control individuals (CTRL), one NHD patient, and one MS subject (normal appearing white matter, NAWM; and active lesions in the white matter, MS ACTIVE) were stained with solochrome cyanine staining (SOLOCY). Original magnification: 4X and 20X; scale bar: 250μm and 25μm, respectively. H) Quantification of LAMP1 or I) PLIN2 signal per Iba1^+^ cell. LAMP1 and PLIN2 intensity: BG: CTRL N=3, n= 18 cells; NHD1^+^ N=1, n= 20; FL: CTRL N=2, n= 17 cells; NHD1^++^ N=1, n= 10; MS N=1, n= 20. *P < 0.05, ****P < 0.0001, analysis was performed between two groups using a Mann-Whitney T test.

We next examined microglia morphology in tissues from two NHD patients, three individuals with no evidence of CNS pathology, and two subjects affected by multiple sclerosis (MS), a prototypic neurological disease characterized by myelin loss (Table S1). NHD is an exceedingly rare disease, and tissue from NHD patient brains that have come to autopsy are limited. Given these limitations, we analyzed the basal ganglia of one NHD subject (named NHD^+^) and the frontal lobe of a second NHD subject (named NHD^++^). Highly ramified cells are designated ‘resting’ microglia and amoeboid cells are called ‘activated’, with a range of intermediate activation states in between [54]. By analyzing the ramification index (a morphological measure of microglia activation), we observed that Iba1^+^ cells from NHD^+^ and NHD^++^ patients exhibited a dystrophic appearance and decreased ramification compared to controls, which corresponds to an activated status (Fig. 6D and E). Microglia in active lesions from MS patients were significantly more rounded and less ramified compared to NHD and CTRL subjects, and thus highly activated (Fig. 6E). This observation confirms that microglia from NHD patients are aberrantly activated and dystrophic, but at an intermediate stage compared to MS patients.

To investigate whether lysosomal defects and lipid metabolism are altered in NHD brains, we performed immunohistochemistry in brain tissues from the same subjects as above (three controls, two NHD patients and two MS patients). We observed that the lysosomal membrane glycoprotein LAMP1 was upregulated in Iba1^+^ microglia in MS tissues compared to controls (Fig. 6F and H). In NHD^+^, LAMP1 intensity was significantly higher in Iba1^+^ cells, however, in NHD^++^, LAMP1 intensity was not changed compared to controls (Fig. 6F and H). This observation is consistent with the dysfunction in the lysosomal machinery and accumulation of degradative vesicles described in NHD iMGLs (Fig. 4). Strikingly, tissues from NHD subjects exhibited areas of diffuse demyelination compared to control subjects, which was similar to white matter (WM) active lesions detected in the MS subjects (Fig. 6G). However, while myelin debris were largely cleared from the tissue in the MS lesion, in NHD, myelin debris remained, as shown by a diffuse blue staining (Fig. 6G). The defect in myelin debris clearance may be due to the reduced degradative capacity of the NHD microglia owing to the observed lysosomal defects. We next stained for PLIN2 in control, MS and NHD brain tissues (Fig. 6F and I). There was a significant increase in PLIN2 signal in Iba1^+^ cells in the MS active lesions; however, despite microglial activation and the inflammation observed in NHD tissues, PLIN2 staining in NHD^+^ and NHD^++^ was significantly lower than the signal detected in MS subjects and was comparable to controls (Fig. 6F and I). This result is consistent with the findings in NHD iMGLs showing reduced lipid droplets, PLIN2, and Bodipy staining in the presence and absence of myelin (Fig. 5). Thus, while demyelination and lipid metabolism are both involved in NHD and MS [13], the cellular phenotypes exist on a spectrum, which may reflect different pathological mechanisms driving these clinically distinct phenotypes.

## DISCUSSION

Here, by combining transcriptomic and functional analyses in control and NHD iMGLs and brain tissues from NHD patients, we further improved our understanding of the mechanisms underlying this rare leukodystrophy. We showed that iMGLs derived from patients affected by NHD and carrying the *TREM2* p.Q33X mutation display dysregulation of lysosomal function, reduced lipid droplets, and downregulation of cholesterol metabolism genes. The defective activation and downregulation of HLA-DR molecules detected in NHD iMGLs were rescued by enhancing lysosomal biogenesis through mTOR-dependent and independent pathways. Alteration in the same pathways were also observed in post-mortem brain tissues from NHD patients, closely recapitulating *in vivo* the phenotype observed in iMGLs *in vitro*.

Thus far, murine models are not able to recapitulate the key phenotypic features observed in patients affected by NHD. This has become an increasingly pressing challenge as rare variants in *TREM2* have been identified as risk factors in Alzheimer’s disease and frontotemporal dementia [9, 35, 36, 38, 50]. Stem cell technologies have allowed us to obtain human cells from clinically defined patients, allowing for in-depth phenotypic analyses in exceedingly rare diseases such as NHD. Therefore, leveraging iMGLs represents a unique modeling platform to define disease-relevant, patient-specific phenotypes in a dish to understand disease mechanisms and for eventual therapeutic target identification, validation and drug discovery pipelines.

Transcriptional and functional analyses highlight that NHD iMGLs *in vitro* are hyporeactive compared to iMGLs from healthy controls, showing a deficit in HLADR, CD80, and CD86 costimulatory molecules expression. This phenotype is reminiscent of what has been described in iMGL-chimeric mouse models harboring *TREM2* deficient microglia [63]. Single cell RNAseq on iMGLs isolated from engrafted mice showed that *TREM2* KO iMGLs were shifted toward a more homeostatic profile even in wild-type mice, with a specific decrease of genes involved in the MHC-II and HLA presentation. Thus, the absence of TREM2 maintains iMGLs in a quiescent state, but this dormant state is released after LPS activation [63]. Conversely, in NHD and controls brain tissues, Nanostring analysis revealed the upregulation of genes involved in immune cell recruitment and activation, inflammation, microgliosis and astrocytosis. The discrepancy between the inflammatory status observed in NHD tissues and the hyporeactive phenotype detected in iMGLs in culture, may be explained by the nature of the model system, where iMGLs capture function and transcriptome of microglia isolated in culture, while post-mortem tissues are an end-stage snapshot resulting from the interaction of microglia with a much more complex cellular milieu after years of accumulating pathology. One possible scenario is that NHD microglia could be hypoactive early during the disease, and this abortive activation in NHD brains could lead later to a dystrophic and pro-inflammatory microglial phenotype. The hyperactivation observed in NHD iMGLs *in vitro* after LPS challenge indeed supports this possible scenario. To further support this hypothesis, in a model of acute CNS demyelination, lack of TREM2 was associated with defective microglia activation, while in chronic demyelination, TREM2 deficient microglia acquired a more pro-inflammatory and potentially neurotoxic phenotype [13].

Multiple groups have examined the impact of NHD mutations on cellular phenotypes in iPSC-iMGLs models. These efforts have focused on the NHD mutations *TREM2* p.*T66M, p.W60C, p.Y38C, p.V126G* [12, 18, 30, 46, 49, 68, 75]. Despite differences in the protocols adopted to generate human microglia, a common phenotype reported includes a reduction in TREM2 and sTREM2 expression, defects in phagocytosis of apoptotic bodies, and reduced survival in the mutant microglia. LPS-mediated cytokine secretion was comparable between control and TREM2 missense mutations in one study [30], while another group came to the opposite conclusion [12]. Microglia-like cells from patients carrying *TREM2* p.T66M, in either homozygous and heterozygous state, or *TREM2* p.W50C mutations, responded to LPS stimulation and were phagocytically competent despite accumulation of an immature form of TREM2 that was not trafficked to the plasma membrane [12]. We observed common gene signatures shared by *TREM2* p.Q33X carriers and *TREM2* deficient iMGLs [63]. These findings suggest that some, but not all, of the observed defects are driven by *TREM2* loss of function. Our sequencing results and functional experiments also highlights the differences between NHD1 and NHD2 iMGLs. This may be due to differences in the clinical severity of disease, other genetic modifiers, and/or sex-based microglial effects. Further studies are required to address these observations. Thus, studies of *TREM2* mutations are essential for capturing the full complexity of disease.

We show that lysosomal dysfunction is a common phenotype shared across NHD iMGLs and NHD brains. NHD iMGLs exhibit reduced acidification and proteolytic capacity of degradative vesicles along with an accumulation of undegraded material in multivesicular bodies. To this end, we hypothesize that the accumulation of unprocessed material within the MVBs observed in NHD iMGLs is a consequence of the lysosomal defects. Similarly, we observed lysosomal defects in NHD brains, as shown by increased LAMP1 protein expression, which marks degradative vesicles, in the basal ganglia and decreased transcription of genes implicated in lysosomal acidification (*ATP6AP2)* and chaperone mediated autophagy (*LAMP2*). The dysregulation of lysosomal genes and accumulation of lysosomal debris have been widely reported in lysosomal storage diseases [81]. Lysosomal dysfunction has broad consequences on cellular function and may affect core signaling events of the cell [5, 8]. In this regard, MHC class II molecules and recycling of peptides displayed by MHC class II molecules are assembled in the endo-lysosomal compartment [17, 28]. Thus, the reduction observed in MHC-I and II genes and proteins in NHD iMGLs might be explained by the deficient endo-lysosomal transport preventing the correct assembly and delivery of these molecules at the membrane. By targeting lysosomal and autophagic processes through Torin1 and curcumin C1 analog, we fully restored HLA-DR levels in NHD iMGLs. This is the first evidence that the lysosomal compartment could be a critical hotspot impacted during the disease by the presence of TREM2 mutations in human microglia, largely responsible for NHD pathophysiology. These findings suggest that the endolysosomal degradative pathway could be a novel and attractive therapeutic target for this disease or others characterized by altered TREM2 function in microglia [25, 81].

*TREM2* p.Q33X iMGLs show downregulation of cholesterol metabolism at the gene network and protein level. We observe a reduced amount of lipid droplets in NHD iMGLs compared to controls, both in the presence or absence of myelin. Lipid droplets are dynamic subcellular organelles required for storage of neutral lipids such as glycero-lipids and cholesteryl esters, and have been recently described as essential to metabolically support immune responses, antigen cross-presentation, interferon (IFN) responses, and production of inflammatory mediators [69]. Alteration in myeloid cell activation status is connected to profound changes in lipid droplets numbers and composition. Lipid droplets have also been shown to accumulate in microglia during aging, being part of a characteristic microglial transcriptional signature called lipid-droplets-accumulating microglia (LDAM), which show defects in phagocytosis, release elevated levels of proinflammatory cytokines and reactive oxygen species [60]. Observations of reduced lipid droplet content in NHD iMGLs are consistent with work showing that, upon demyelinating injury, *Trem2*-deficient mice are unable to elicit the adaptive response to excess cholesterol exposure, form fewer lipid droplets than wild-type mice, and develop endoplasmic reticulum stress [32]. This supports the hypothesis that the TREM2 receptor is required for lipid droplets biogenesis and to buffer cholesterol-mediated toxicity in mouse and human microglia. Another possible interpretation of our findings, is that lysosomal dysfunction in NHD iMGLs would lead to an altered degradation of these lipidic structures [96].

Finally, we observed the downregulation of genes involved in neuroplasticity, synaptic function and microglia-to-neuron cross-talk in NHD brains versus controls. Microglial survival is controlled by IL34 and colony-stimulating factor 1 (CSF1), through CSF1R signaling activation [22, 39]. Microglia, in turn, are critical for the regulation of neuronal activity and firing in a region-specific and microglia number-dependent manner [4]. This highlighted by observations that NHD patients do not only display defects at the microglial level but instead present with neuronal phenotypes and dysfunction that might be microglial-dependent and/or independent. This area of study raises many new questions about the impact of TREM2 mutations on neuronal function during neurodegeneration and highlights how studying NHD might help us understand also the microglia-to-neuron cross-talk and the impact of TREM2 mutations on this interaction. Overall, our results suggest that NHD might be considered a lysosomal-storage disease with white matter alterations as first proposed in reports based solely on NHD pathology [45], when the genetic cause of the disease was not known. Our study provides the first cellular and molecular evidence that lack of TREM2 in microglia leads to a defect in lysosomal function. A better understanding of how microglial lipid metabolism and lysosomal machinery are altered in NHD may provide new insights into mechanisms underlying NHD pathogenesis.

## MATERIALS AND METHODS

### Patient Consent

Skin biopsies were collected following written informed consent from the control unaffected sibling or the parents for the NHD affected individuals. The informed consent was approved by the Washington University School of Medicine Institutional Review Board and Ethics Committee (IRB 201104178).

### Human postmortem brain tissue

Human tissue sections were obtained from two NHD patients, three control subjects and two MS patients. The two MS patients were obtained from The Neuroinflammatory Disease Tissue Repository at Washington University St. Louis. Brain autopsies from NHD tissues were obtained from the University of Washington Neuropathology Core Brain Bank and the Meiji Pharmaceutical University, Tokyo, Japan. Under protocols approved by the Institutional Review Boards of the University of Washington and the Meiji Pharmaceutical University, all subjects had previously given informed consent to share and study autopsy material. All methods for processing and analyzing the brain autopsy tissues followed relevant guidelines and regulations. Demographic and clinical characteristics of the donors of human brain tissues at the time of collection are indicated in Table S1.

### Dermal fibroblast isolation

Dermal fibroblasts were isolated from skin biopsies obtained from research participants in an NHD family. Briefly, skin biopsies were collected by surgical punch and stored in Fibroblast Growth Media (Lonza). To isolate dermal fibroblasts from skin biopsy, the biopsies were rinsed with PBS and cut lengthwise with dissecting scissors. The resulting tissue sections were then plated into a dry 24-well tissue culture treated plate (approximately 6-12 sections). After removing excess PBS from the wells, 300µL of fibroblast growth media (Lonza) was carefully added and tissue was incubated at 37°C and 5% CO_2_. After 24 h, tissue was supplemented with 1mL fibroblast growth media and media changes were repeated every 3-4 days. Fibroblast cells were observed to migrate from the tissue within 2 weeks of culture. Dermal fibroblasts were maintained in Fibroblast Growth Media (Lonza) supplemented with penicillin/streptomycin.

### Macrophage differentiation from PMBCs

Peripheral blood mononuclear cells (PBMCs) were purified from human blood on Ficoll-Paque PLUS density gradient (Amersham Biosciences, Piscataway, NJ). To generate macrophages, PBMCs were cultured in 6-well culture plates (3×10^6^ cell/well) in RPMI-1640 without fetal bovine serum. After 2 hours of culture, PBMCs were washed twice with PBS 1X and cultured in RPMI supplemented with 50 ng/mL MCSF for 7 days at 37° C with 5% CO_2_.

### iPSC generation and characterization

Human fibroblasts (IDE: SB CTRL, F12455: NR CTRL, IDO: NHD1, and 1F1: NHD2) were transduced with non-integrating Sendai virus carrying the four factors required for reprogramming into iPSC: OCT3/4, SOX2, KLF4, and cMYC [6, 89]. Cells showing morphological evidence of reprogramming were selected by manual dissection. Human iPSCs were cultured using feeder-free conditions (Matrigel, BD Biosciences, Franklin Lakes, NJ, USA). Human iPSCs were thawed (1-2 x 10^6^ cells/mL), diluted in DMEM/F12, and centrifuged at 750 rpm for 3 minutes. The resulting iPSC pellet was then diluted in mTeSR1 supplemented with Rock inhibitor (Y-27632; 10µM final). iPSCs were subsequently cultured in 37°C, 6% CO_2_ with daily medium changes (mTesR1, STEMCELL Technologies, Vancouver, BC, CA). All iPSC lines were characterized using standard methods [89]. Each line was analyzed for chromosomal abnormalities by karyotyping (Figure S1A), for pluripotency markers (OCT4A, SOX2, SSEA4, TRA1) by immunocytochemistry (ICC) (Invitrogen A24881) (Fig. S1B), and for *TREM2* mutation status (homozygous p.Q33X mutation) by Sanger sequencing (Fig. S1C).

### iPSC-derived microglia-like cells

iMGLs were generated as previously described in [1, 24, 62]. iPSCs were differentiated into hematopoietic precursors cells (HPCs) using a STEMdiff Hematopoietic kit (STEMCELL Technologies) and following manufacturer’s instructions. Briefly, to begin HPC differentiation, iPSCs were detached with ReLeSR™ (STEMCELL Technologies) and passaged in mTeSR1 supplemented with Rock inhibitor to achieve a density of 100-200 aggregates/well (for iPSCs from mutation carrying donors, aggregate numbers to be plated requires line-specific optimization). On day 0, cells were transferred to Medium A from the STEMdiff Hematopoietic Kit. On day 3, flattened endothelial cell colonies were exposed to Medium B and cells remained in Medium B for 10 additional days while HPCs began to lift off the colonies. After 12 days in culture, HPCs were collected by removing the floating population with a serological pipette and the adherent portion after incubation with Accutase™ (STEMCELL Technologies) for 15 min at 37°C. The floating and adherent CD43^+^ population (see below *Fluorescent Activated Cell Sorting (FACS)* section) was sorted with a Becton Dickinson FACSAria II cell sorter. At this point, HPCs can be frozen in CryoStor® CS10 (STEMCELL Technologies) and stored in liquid nitrogen. iMGL induction was achieved by culturing CD43^+^ HPCs in iPSC-Microglia medium (DMEM/F12, 2X insulin-transferrin-selenite, 2X B27, 0.5X N2, 1X glutamax, 1X non-essential amino acids, 400 mM monothioglycerol, and 5 mg/mL human insulin) freshly supplemented with 100 ng/mL IL-34, 50 ng/mL TGFβ1, and 25 ng/mL M-CSF (Peprotech) for 25 days (37 days from iPSC). During the last 3 days in culture, 100 ng/mL CD200 (Novoprotein) and 100 ng/mL CX3CL1 (Peprotech) were added to iPSC-Microglia medium to mimic a brain-like environment (Supplementary Fig. 2A). All the subsequent analyses described in the paper have been performed on fully mature iMGL (between day 40 to day 52) unless otherwise stated.

### Fluorescent Activated Cell Sorting (FACS)

All steps were performed on ice or using a pre-chilled refrigerated centrifuge set to 4°C with all buffers/solutions pre-chilled before addition to samples. For HPC sorting, HPCs were collected using sterile filtered FACS buffer (1X DPBS, 2% BSA), cells were then filtered through 70 μm filters to remove large clumps, washed with FACS buffer (300 x g for 5 min 18□C), then stained for 20 min at 4°C in the dark using the following antibodies: CD43-APC (Cat#: 343206, Clone: 10G7), CD34-FITC (Cat#: 343504, Clone: 581), CD45-Alexa 700 (Cat#: 304024; Clone: H130) from BioLegend, and Zombie Aqua™ Fixable Viability Kit (BioLegend). The CD43^+^ total population was sorted with a Becton Dickinson FACSAria II cell sorter. For detection of microglial surface markers, iMGLs were incubated for 10 min on ice with anti-CD16/CD32 to block Fc receptors (1:50; Miltenyi Biotec, Cat #:120-000-442) and with Zombie Aqua™ to identify live cells. Then, iMGLs were stained with CD11b-PeCy7 (Cat#: 101216; Clone: M1/70), CD45-Alexa 700 (Cat#: 304024; Clone: H130), CD80-BV421 (Cat#: 305222, Clone: 2D10), CD86-PerCP/Cy5.5 (Cat#: 30420, Clone: IT2.2) all from BioLegend, CD14-PeCy7 (Invitrogen, Cat#: 25-0149-41, Clone: 61D3), HLA-DR-PECF594 (BD, Cat#: 562331 clone: G46-6), TREM2-APC (R&D, FAB17291A), TREM2-biotinilated (Clone: E29E3, generously provided by Dr. Marco Colonna) for 20 min at 4°C in the dark. AnnixinV/Propidium iodide positive cells were detected with a FITC Annexin V Apoptosis Detection Kit with PI (Biolegend). Cells were acquired on a BD LSRFortessa and BDX20 and data analyzed with FlowJo software (FlowJo).

### Lysosomal acidity measurement

To evaluate lysosomal acidity, iMGLs were incubated with 5nM of LysoTracker® Red DND-99 (ThermoFisher, L7528), diluted in the cell medium at 37°C for 5 min. Live cell images were acquired with a Nikon Eclipse 80i fluorescent microscope and Metamorph Molecular Devices software. The fluorescence intensity of LysoTracker Red staining in the soma was quantified by Fiji. Briefly, the fluorescence intensity of LysoTracker® in the soma of each cell was measured and then corrected for background fluorescence resulting in the Corrected Total Cell Fluorescence (CTCF) values. To study the overall protease activity, iMGLs were incubated for 4 h at 37°C with 10µg/mL of DQ™ Red BSA (ThermoFisher, D12051) diluted in the cell medium. Cells were washed once with HBSS solution. Live cell images were acquired immediately with a Nikon Eclipse 80i fluorescent microscope and Metamorph Molecular Devices software. Fluorescence intensity of DQ™ Red BSA staining in the soma was quantified by Fiji as previously described [61]. DQ™ Red BSA stained cells were next fixed for further immunofluorescence analyses.

### Myelin production and iMGL treatment

Human myelin was prepared as previously described [15, 66] and stored in lyophilized form at −80°C. Prior to use, myelin was suspended in DMEM to a final concentration of 2 mg/mL and dissolved by vortexing and sonicating. Myelin was then irradiated with 10,000 RADS to achieve sterility. Aliquots were stored at −80°C for further use. For treatment with myelin, iMGLs were plated on Matrigel-coated 12-mm coverslips at 5 × 10^4^ cell/well and incubated with 10 μg/mL of pHrodo-labeled myelin (pHrodo™ Red, SE, ThermoFisher) for 24h.

### Torin1 and curcumin analog C1 treatment in iMGL

Fully mature iMGLs were plated in 24-well plate at a density of 1 × 10^5^ cell/well and treated with 250 nM Torin1 (Tocris, 4247) for 4 h or with 1μM of curcumin analog C1 (Cayman Chemical Company, 34255) for 7 h or with DMSO as control. After 4 h and 7 h of treatment respectively, iMGLs were washed once in PBS and cultured in complete iPSC-Microglia medium for 24 h. Cells were then collected and stained for detection of microglial surface markers by FACS.

### Immunocytochemistry

Cells were washed three times with DPBS (1X) and fixed with cold PFA (4% w/v) for 20 min at room temperature (RT) followed by three washes with PBS (1X). Cells were blocked with blocking solution (PBS with 0.1% Triton X-100 and 1% BSA) for 30 min at RT. Primary antibodies were added at respective dilutions (see below) in blocking solution and placed at 4°C overnight. The next day, cells were washed 3 times with PBS for 5 min and stained with Alexa Fluor conjugated secondary antibodies from Invitrogen (1:800) for 2.5 h at room temperature in the dark. After secondary staining, cells were washed 3 times with PBS and cover slipped with ProLong™ Diamond Antifade Mountant (ThermoFisher). Primary antibodies used for ICC: anti-P2ry12 (1:500, HPA014518 Sigma), anti-TREM2 (1:200, AF1828 R&D Systems), anti-CD68 (1:100, MO718 Dako), anti-TMEM119 (1:100, ab185333 Abcam), anti-IBA1 (1:500; Wako, 019-19741). *Lipid droplets staining:* for Perilipin 2 (PLIN2) staining, cells were blocked in PBS with 0.1% saponin and 1% BSA for 5 min and subsequently incubated with ADRP/Perilipin 2 antibody (1:500, 15294-1-AP Proteintech) in PBS with 0.1% saponin and 1% BSA overnight at 4°C. For BODIPY staining, cells were incubated in PBS with BODIPY FL Dye (1:1000 from a 1 mg/mL stock solution in DMSO; ThermoFisher) a dye that specifically labels neutral lipids [83] and Hoechst 33342 (1:5000; ThermoFisher) for 20 min at room temperature (RT). To analyze the percentage of lipid-droplet-containing iMGLs, the numbers of total Hoechst+ cells and of Hoechst^+^ cells with BODIPY^+^ lipid droplets were counted, and the percentage of BODIPY^+^ iMGLs was calculated. For confocal analysis, images were acquired with a Zeiss LSM980 Airyscan 2 laser scanning confocal microscope (Carl Zeiss Inc., Thornwood, NY) equipped with 63X and 40X, 1.4 numerical aperture (NA) Zeiss Plan Apochromat oil objectives. The system was equipped with a unique scan head, incorporating a high-resolution galvo scanner along with two PMTs and a 32-element spectral detector. ZEN 3.4.9 Blue edition software was used to obtain Z-stacks through the entire height of the cells. Images taken were optimized for 1 airy unit using the 405 nm diode, 488 nm diode, and 561 nm diode and 633 nm HeNe (helium neon) lasers. Images were finally processed with ImageJ and Imaris Software (Bitplane, Switzerland). Quantification was performed on original orthogonal Z-projections generated in ImageJ software. The particle numbers were quantified with ImageJ 1.5j8 (NIH) with size (pixel2) settings from 0.1 to 10 and circularity from 0 to 1. PLIN2-positive droplets with intensity above cytoplasmic background and size (pixel2) from 10 were quantified. A total of 40-50 cells per line were analyzed.

### Histology and immunohistochemistry of formalin-fixed human brain tissue

Formalin-fixed paraffin embedded (FFPE) 5 μm human tissue sections were stained with solochrome cyanine to detect area of demyelination. For immunohistochemistry analysis, sections were deparaffinized and rehydrated in xylene, then sequential concentrations of ethanol to water followed by antigen retrieval in boiling 0.01M citric acid pH 6.0 for 10 min. Sections were then incubated in PBS with 5% horse serum and 0.1% triton-X for 1 h at RT followed by incubation overnight at 4°C with primary antibodies: goat anti-Iba1 (1:250; Novus, NB100-1028), rabbit anti-PLIN2 (1:100; 15294-1-AP, Proteintech) and monoclonal LAMP1 (H4A3, undiluted, developed by Developmental Studies Hybridoma Bank). After primary antibodies incubation, sections were washed three times in PBS and incubated in PBS with Alexa Fluor conjugated secondary antibodies from Invitrogen (1:1000) for 1 h at RT in the dark. Sections were mounted with Vectashield (Vector Laboratories, H-1000). Quantitative evaluation of microglial cell morphology in tissue sections was performed as described in the literature using ramification index (RI) calculated by the following equation: 4π × cell area/(cell perimeter)^2^ [13]. The RI of perfectly round cells is 1; if morphology deviates from circular form, RI is smaller than 1; when the cell is highly ramified the RI is close to zero. The images were acquired with a Nikon Eclipse Ni fluorescent and bright field microscope equipped with 10X, 20X, and 60X zoom objectives. Cell area and perimeter of Iba1^+^ cells and LAMP1^+^ or PLIN2^+^ signal per Iba1^+^ cell were calculated with the assistance of NIS-Elements software.

### sTREM2 ELISA

A sandwich ELISA for human soluble TREM2 (sTREM2) was developed as previously described [20]. Briefly, an anti-human TREM2 monoclonal antibody (R&D Systems, catalog no. MAB1828, clone 263602; 0.5 mg/ml) was used as a capture antibody and coated overnight at 4°C on MaxiSorp 96-well plates (Nalgene Nunc International, Rochester, NY) in sodium bicarbonate coating buffer (0.015 M Na2CO3 and 0.035 M NaHCO3, pH 9.6). Washes between the steps were done four times with PBS/0.05% Tween 20 (Sigma-Aldrich). Wells were then blocked for 4 h at 37°C with 10% fetal bovine serum (FBS) in phosphate-buffered saline (PBS). Freshly thawed supernatants and recombinant human TREM2 standard (SinoBiological, catalog no. 11084-H08H-50) were incubated in duplicate overnight at 4°C. For detection, a goat anti-human TREM2 biotinylated polyclonal antibody (R&D Systems, catalog no. BAF1828; 0.2 mg/mL) was diluted in assay buffer (PBS/10% FBS at 1:3000) and incubated for 1.25 h at RT on an orbital shaker. After washing, wells were incubated with horseradish peroxidase–labeled streptavidin (BD Biosciences, San Jose, CA; diluted 1:3000) for 1 h at RT with orbital shaking. Horseradish peroxidase visualization was performed with 3,3′,5,5′ tetramethylbenzidine (Sigma-Aldrich, St. Louis, MO) added to each well for 10 min at RT in the dark. Color development was stopped by adding an equal volume of 2.5 N H2SO4. Optical density of each well was determined at 450 nm. Samples were run in duplicate in each assay. Raw values are provided as [pg/ml].

### RNA extraction, sequencing, and transcript quantification

Total RNA was isolated from iMGLs cells using the RNeasy Mini Kit (Qiagen, Hilden, Germany) according to the manufacturer’s protocol. For iMGLs cell pellets frozen in Qiazol, 200µL of chloroform was added, then the samples were shaken for 5 seconds and centrifuged at 12,000g for 15 min or until complete phase separation. 400-600µL aqueous phase was then transferred into a new tube and cleaned using reagents and spin columns from the RNeasy Mini Kit. Cell line samples were DNase-treated using reagents from the RNase-Free DNase Set (Qiagen, Hilden, Germany). TapeStation 4200 System (Agilent Technologies) was used to perform quality control of the RNA concentration, purity, and degradation based on the estimated RNA integrity Number (RIN), and DV200. Extracted RNA (10µg) was converted to cDNA by PCR using the High-Capacity cDNA Reverse Transcriptase kit (Life Technologies). Samples were sequenced by an Illumina HiSeq 4000 Systems Technology with a read length of 1×150 bp, and an average library size (mapped reads) of 36.5 ± 12.2 million reads per sample.

Identity-by-Descent (IDB) [11] and FastQC (Andrews *et al.*, 2012) analyses were performed to confirm sample identity. STAR (v.2.6.0) [21] was used to align the RNA sequences to the human reference genome: GRCh38.p13 (hg38). The quality of RNA reads including the percentage of mapped reads and length of sequence fragments was performed. Aligned sequences were processed using SAMtools (v.1.9) [55]. The average percentage of unique mapped reads in the BAM files was 82.5% ± 3.62, and the average percentage of mapped reads to GRCh38 was 89.77% ± 5.12. Salmon (v. 0.11.3) [76] was used to quantify the expression of the genes annotated. Protein coding genes (a total of 60754 genes) were selected for further analyses (Table S2).

### Principal component analyses and differential expression analyses

Human macrophages and iMGLs were plotted using the plotly package [56] based on regularized-logarithm transformation (rlog) counts and utilizing the 500 most variable protein coding genes. Differential gene expression between WT and NHD carriers was calculated using the DESeq2 (v.1.22.2) package [58]. The transcripts per million (TPM) of the gene set of interest were made into heatmaps with ComplexHeatmap [34]. In iMGLs, the TPM was further normalized by the expression of *GAPDH*. The top 500 genes were made into a PCA graph to investigate the differences between samples. The DEG analyses compare the expression between WT and NHD carriers in different cell types and were further made into a volcano plot with ggplot2. The significant DEGs (FDR<0.05, unless otherwise stated) were split into upregulated and downregulated DEG, and separately searched for pathway enrichment by EnrichR [14, 52, 94].

### RNA expression analysis by NanoString nCounter

RNA was isolated from FFPE sections, as previously described [50]. Briefly, for each sample, four 10 micron sections were processed using the Recover All RNA isolation kit (Ambion, ThermoFisher). RNA yield and fragment-length distribution were measured by Qubit Fluorimeter (Molecular Probes, ThermoFisher) and 2100 Bioanalyzer (Agilent Technologies). 1 μg of total RNA was hybridized with the PanCancer Immune panel (NanoString Technologies). Counts were normalized and log2 transformed using the nSolver 3.0 Analysis Software (NanoString Technologies).

### Transmission electron microscopy

For ultrastructural analyses, iMGLs were fixed in 2% paraformaldehyde/2.5% glutaraldehyde (Ted Pella Inc., Redding, CA) in 100 mM sodium cacodylate buffer, pH 7.2 for 2 h at RT. Samples were washed in sodium cacodylate buffer and postfixed in 2% osmium tetroxide (Ted Pella Inc) for 1 h at RT. After rinsing extensively in dH_2_O, samples were then bloc stained with 1% aqueous uranyl acetate (Electron Microscopy Sciences, Hatfield, PA). Samples were washed in dH_2_O, dehydrated in a graded series of ethanol, and embedded in Eponate 12 resin (Ted Pella Inc). Sections of 95 nm were cut with a Leica Ultracut UCT ultramicrotome (Leica Microsystems Inc., Bannockburn, IL), stained with uranyl acetate and lead citrate, and viewed on a JEOL 1200 EX transmission electron microscope (JEOL USA Inc., Peabody, MA) equipped with an AMT 8 megapixel digital camera and AMT Image Capture Engine V602 software (Advanced Microscopy Techniques, Woburn, MA). MVB and lipid droplet analyses were performed with ImageJ (Fiji) and the number of MVB and lipid droplets was normalized on the cell area.

## Data Availability

All data produced in the present study are available upon reasonable request to the authors.

## ACKNOWLEGEMENTS

We are thankful to NHD patients and their families for their participation in this research project. We thank Dr. Marco Colonna for providing TREM2 antibody. This work was funded by Centene (CMK), NIH U01 AG052411 (AG), R01 AG058501 (LP), R01 AG062734 (CMK), P30 AG066444 (JM, CMK), UL1 TR002345, and the Chan Zuckerberg Initiative; R01 NS069719 (WHR), the Takayma family gift (OK), the US Department of Veterans Affairs Clinical Sciences R&D Service (Merit Review Award Number 101 CX001702 to WHR and OK). Access to equipment was made possible by the Hope Center for Neurological Disorders, and the Departments of Neurology and Psychiatry at Washington University School of Medicine. Fabia Filipello was supported by the McDonnell Center for Cellular and Molecular Neurobiology fellowship (Washington University in St Louis) and is the recipient of a 2018-0364-CARIPLO grant. Confocal was generated on a Zeiss LSM 880 Airyscan Confocal Microscope which was purchased with support from the Office of Research Infrastructure Programs (ORIP), a part of the NIH Office of the Director under grant OD021629. The recruitment and clinical characterization of research participants at Washington University were supported by NIH P50 AG05681, P01 AG03991, and P01 AG026276. Images created with BioRender.com.

## Supplemental Figure Legends

**Supplemental Figure 1:**
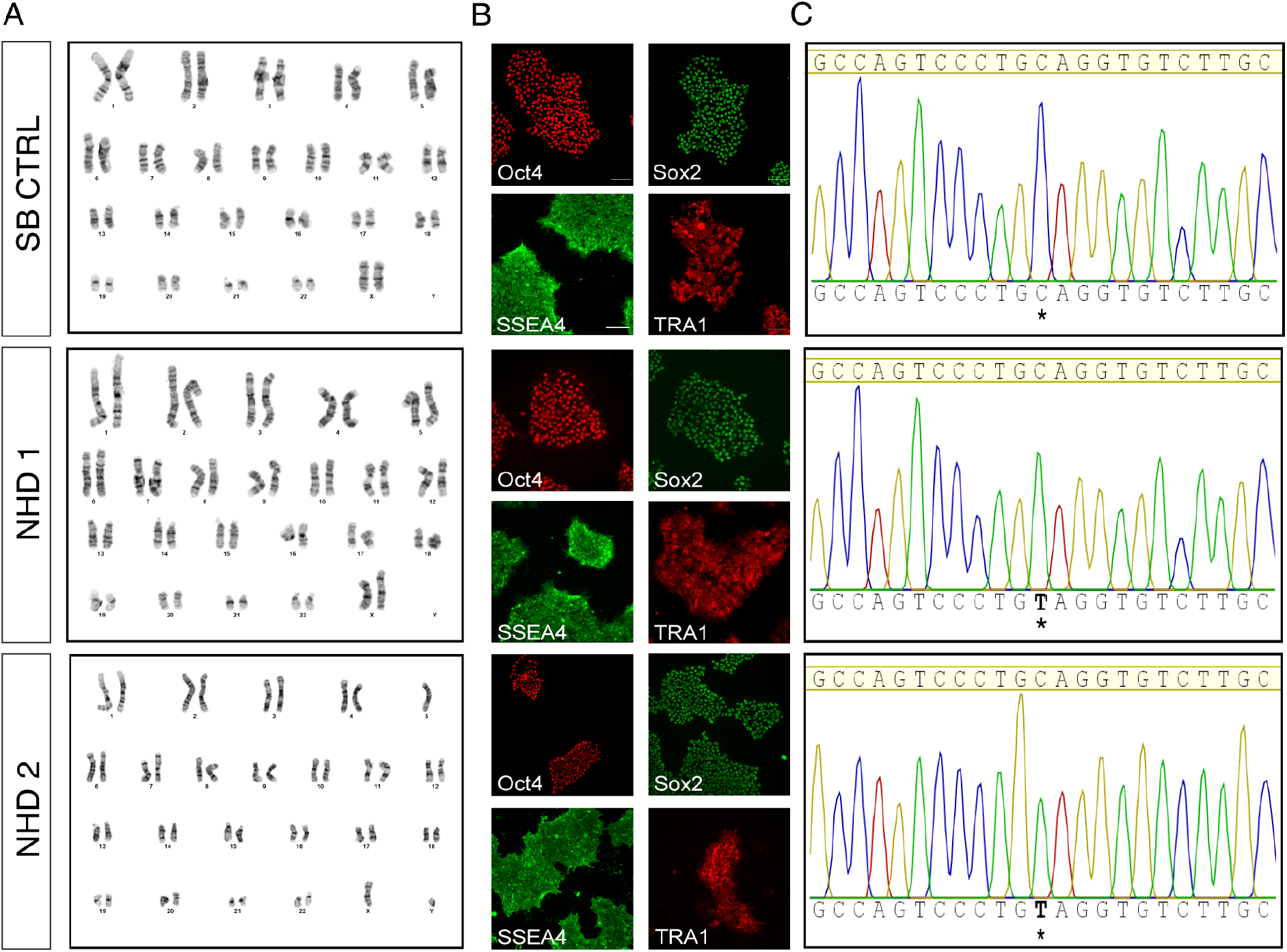
Generation and characterization of iPSCs from NHD patients and non-carrier family member. A) iPSCs from the non-carrier SB CTRL, and the two affected siblings (NHD1 and NHD2) exhibit normal karyotype. B) iPSCs from SB CTRL, NHD1 and NHD2 were stained for octamer-binding transcription factor (OCT4), SRY-Box Transcription Factor 2 (SOX2), Stage-specific embryonic antigen-4 (SSEA4) or Transcription-associated protein 1-60 (Tra-1-60) as indicated. Scale bar 20 μm. C) Sanger sequencing of *TREM2* p.Q33X mutation. Sequence chromatogram showing the C to T point mutation at position 97 (C97T) in exon 2 of the *TREM2* gene. *, variant site. Characterization of unrelated control iPSCs line was reported in [41].

**Supplemental Figure 2:**
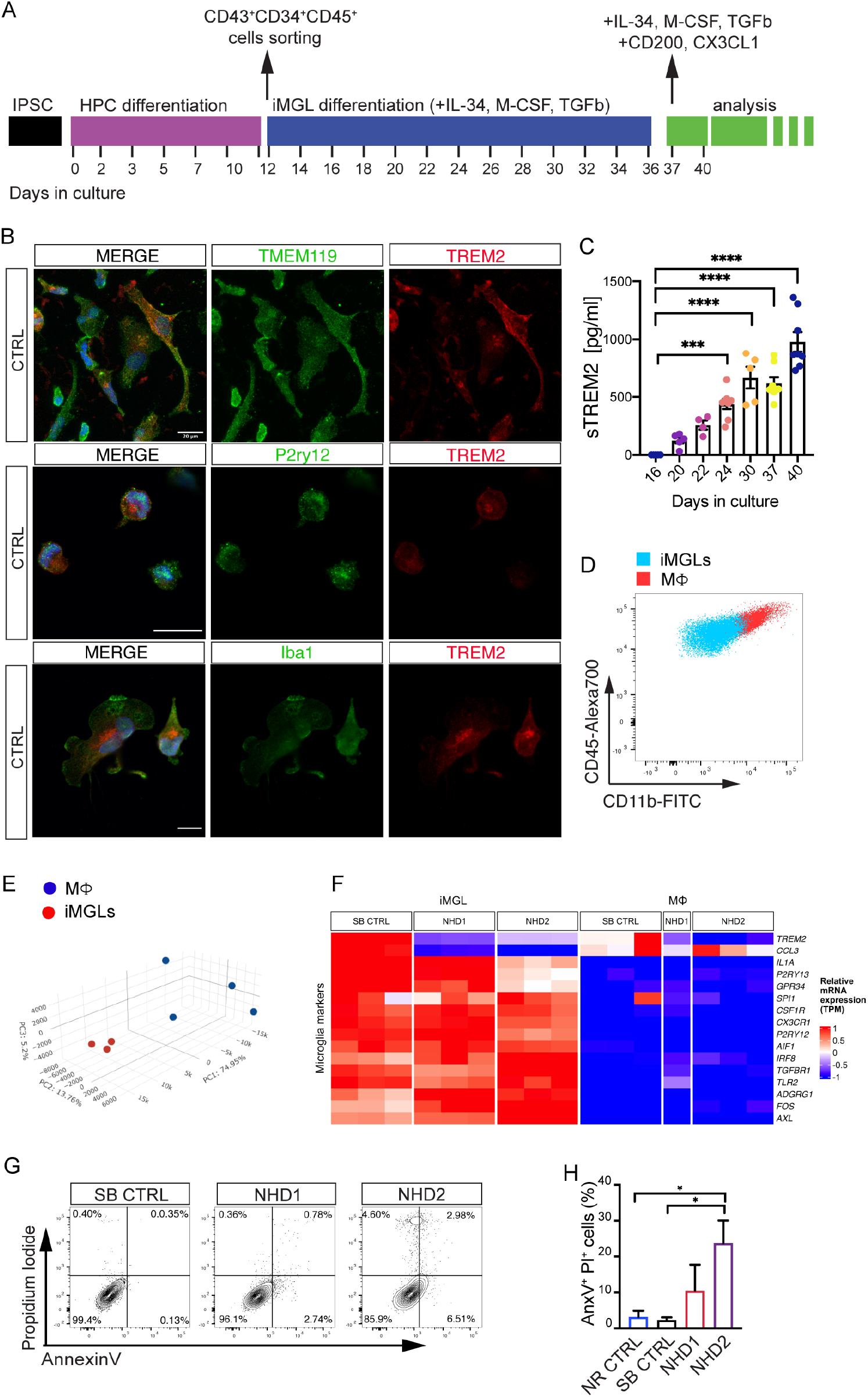
Validation and characterization of fully differentiated iMGLs. A) Schematic of iMGLs differentiation protocol (see Methods for additional information). After 40 days in culture (starting from iPSCs) iMGLs are fully mature and ready for analyses. B) Confocal images of iMGLs from healthy donors (NR CTRL and SB CTRL lines) stained for TMEM119, P2RY12, IBA1 and TREM2. Scale bar, 20 μm. C) sTREM2 detected by ELISA in the media of CTRL iMGLs during the differentiation process. ***P < 0.001, ****P < 0.0001, One-way ANOVA with Dunnett’s post hoc test. Data are pooled from two independent experiments. D) Representative dot plot showing CD11b and CD45 cell surface expression in fully mature iMGLs (blue) versus macrophages (MΦ) from control donors (red). E-F) RNAseq was performed on iMGLs (N=1 SB CTRL, N=2 NHD lines -NHD1 and NHD2-; n=3 replicates/line) and MΦ (N=1 SB CTRL, N=2 NHD lines -NHD1 and NHD2-; n=3 replicates/line for SB CTRL and NHD2, n=1 replicate/line for NHD1) (See Table S1 for donors details). E) 3D principal component analysis (PCA) of iMGLs (red) and MΦ (blue) using the 500 genes with the highest variance (TPM normalized counts). F) Heatmap showing homeostatic microglia signature genes highly expressed by iMGLs compared to M MΦ in SB CTRL versus NHD1 and NHD2 subjects. G) Representative dot plots of SB CTRL, NHD1 and NHD2 iMGLs stained for AnxV and PI and H) relative quantification of absolute AnxV^+^ PI^+^ cells percentage. *P < 0.5, One-way ANOVA with Kruskal Wallis test and Dunn’s multiple comparison test. Data are pooled from three independent experiments. Data shown are mean ± SEM.

**Supplemental Figure 3:**
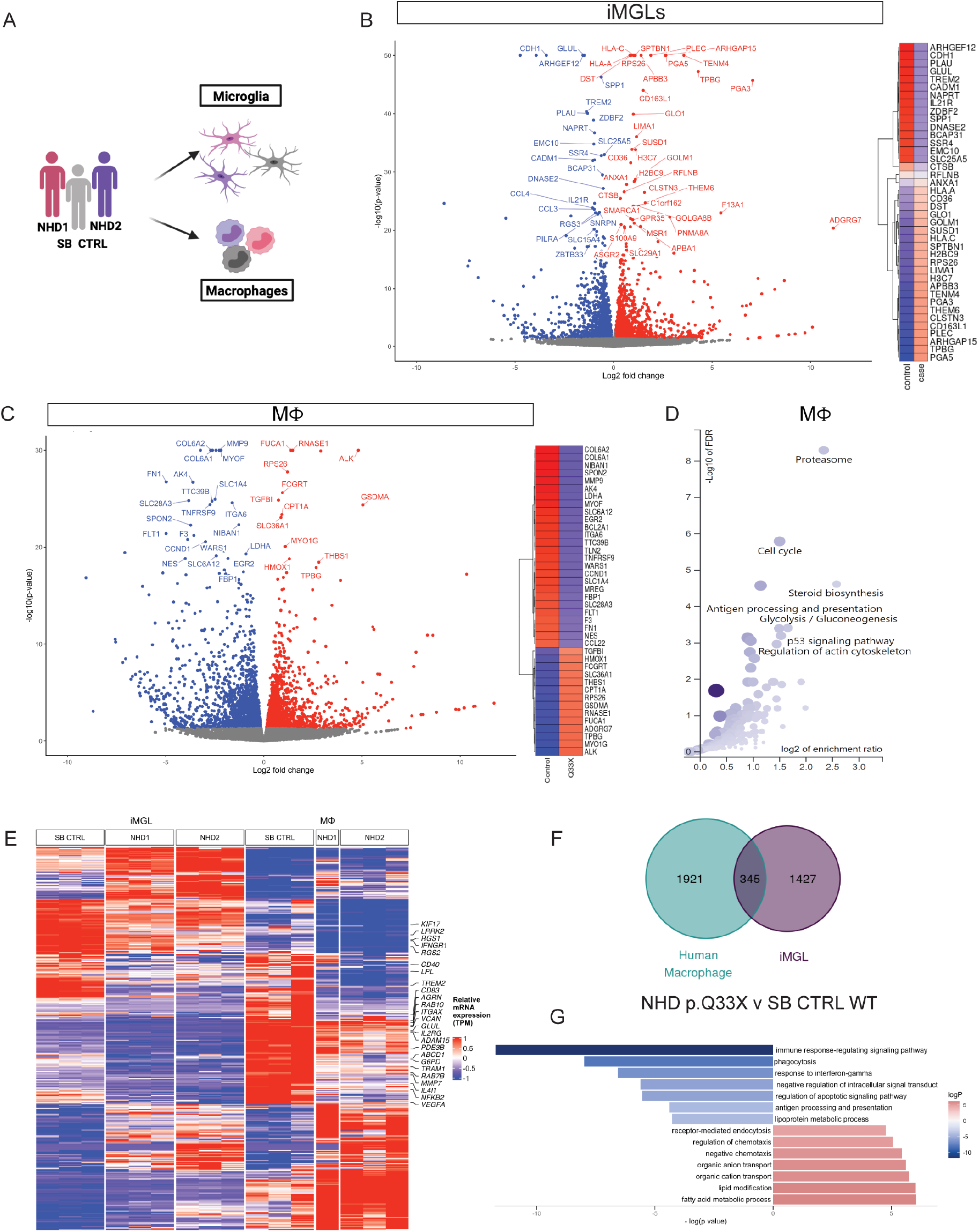
Common pathways shared by NHD iMGLs and human macrophages. A) Cartoon representing iPSCs-derived iMGLs and PBMCs-derived MΦ generation from SB CTRL and NHD donors. B) NHD vs SB CTRL volcano plot of differential gene expression among iMGLs. On the right, are the top significant DEG selected from DEseq2 results. The relative high-low expression levels are represented in red-blue color scale. C) NHD vs SB CTRL volcano plot of differential gene expression among MΦ. On the right, are the top significant DEG selected from DESeq2 results. The relative high-low expression levels are represented in red-blue color scale. D) Pathway analysis of MΦ with down-regulated protein coding DEG (DEG are defined by FDR < 0.05; DEG in Table S4). E) Heatmap of DEG for iMGLs and MΦ from SB CTRL, NHD1 and NHD2. F) Venn diagram of DEG (NHD *versus* SB CTRL) in common between iMGLs and PBMCMΦ (n=345) and G) pathways analysis of DEG in common between iMGLs and MΦ.

**Supplemental Figure 4:**
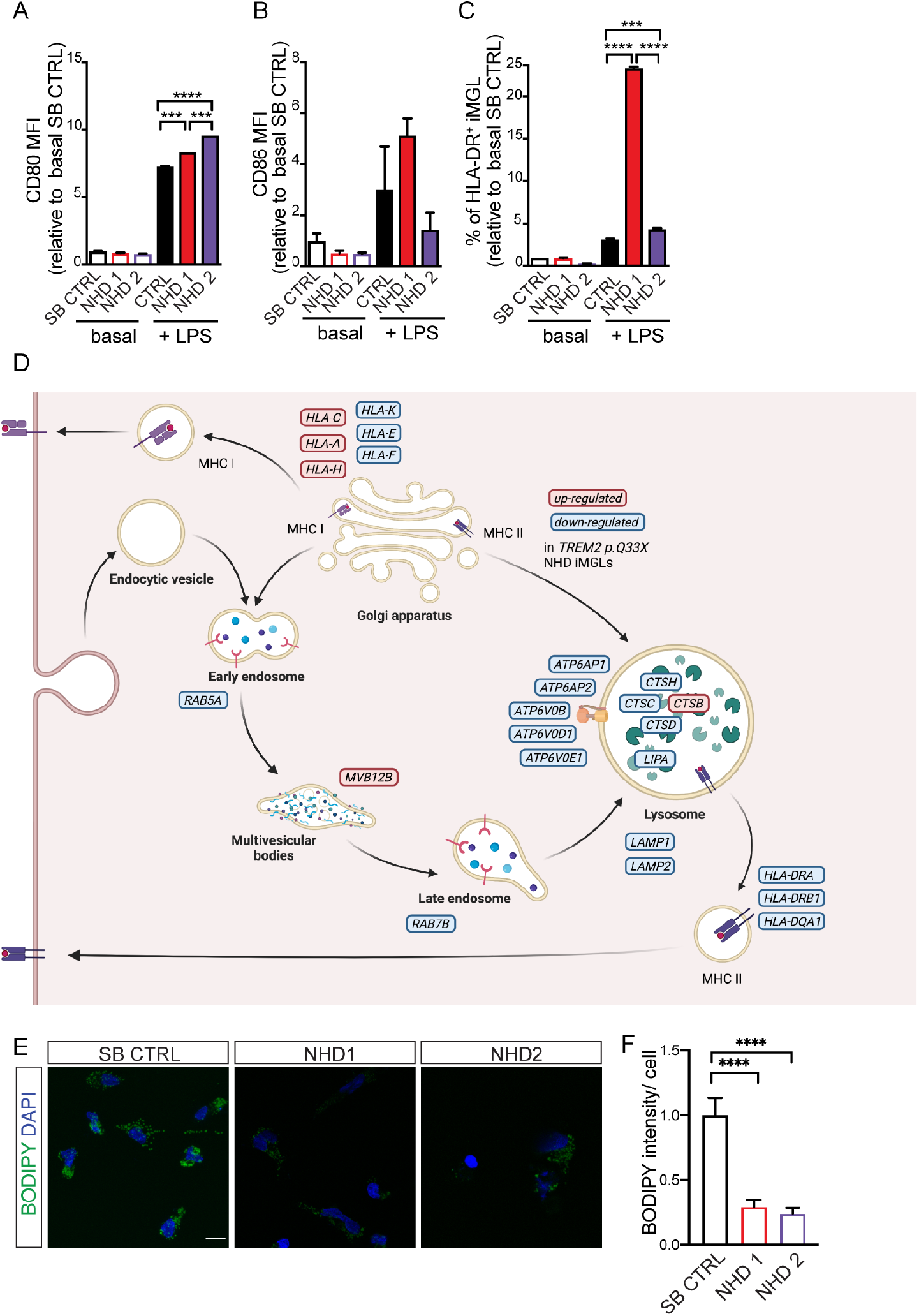
LPS triggers an exaggerated inflammatory response in NHD iMGLs. A-C) FACS analysis of iMGLs from SB CTRL, NHD1 and NHD2 treated for 24 h with LPS (100 ng/ml) and compared to untreated iMGLs. Graphs showing the quantification of surface A) CD80, B) CD86 MFI and C) the percentage of HLA-DR positive cells. ***P < 0.001, ****P < 0.0001, One-way ANOVA with Tukey’s post hoc test. Data are pooled from two independent experiments. D) Cartoon representing the upregulated and downregulated genes in NHD iMGLs compared to SB CTRL belonging to lysosomal and phagocytic pathways. Adapted from BioRender (2022). Endocytic Pathway Comparison (Layout). Retrieved from https://app.biorender.com/biorender-templates/t-61fd5292e07d3d00a5bd4c21-endocytic pathway-comparison-layout. F) Representative images of Bodipy (green), DAPI (blue) staining in iMGLs. Scale bar 10 μm. G) Relative quantification of Bodipy intensity. NR CTRL n= 24 cells; SB CTRL= 26 cells, NHD1 n=30 cells; NHD2 n=33 cells. *P < 0.05, ***P < 0.001, ****P < 0.0001, One-way ANOVA with Kruskal Wallis test and Dunn’s multiple comparison test. Data are pooled from two independent experiments.

